# Open-Label Placebos for Antidepressant Discontinuation Symptoms: A Series of N-of-1 Trials

**DOI:** 10.1101/2025.09.12.25335628

**Authors:** Amke Müller, Stefan Konigorski, Tahmine Fadai, Claire V. Warren, Lena Koschik, Wei Liu, Ulrike Bingel, Winfried Rief, Irina Falkenberg, Tilo Kircher, Yvonne Nestoriuc

## Abstract

**Key points:** **Question** Can open-label placebos (OLP) reduce antidepressant discontinuation symptoms in patients with remitted major depression?

**Findings** This series of N-of-1 trials in 25 patients, over 8 weeks following antidepressant discontinuation, found small effects of OLP vs no treatment in reducing discontinuation symptoms. While the overall treatment effect did not reach the threshold of clinical meaningfulness, 38% of patients experienced a small benefit. Discontinuation symptoms were mostly mild, with few exceptions, and diminished over time.

**Meaning** These findings support the use of personalized approaches and suggest that open-label placebos may be beneficial for antidepressant discontinuation symptoms in some patients.

**Importance:** Antidepressant discontinuation symptoms are highly prevalent and can be severe. Interventions leveraging the placebo effect have the potential to alleviate symptoms and facilitate the discontinuation process.

**Objective:** To evaluate the efficacy of open-label placebo (OLP) in reducing antidepressant discontinuation symptoms.

**Design, Setting, And Participants:** A series of randomized, single-blinded, N-of-1 trials of OLP vs no treatment in an alternating order (ABAB, BABA). Patients with remitted major depressive disorder, reporting moderate to severe discontinuation symptoms after successful antidepressant discontinuation with state-of-the-art clinical supervision were enrolled between December 2021 to December 2023.

**Interventions:** The OLP treatment consisted of a standardized verbal and written rationale and twice-daily placebo intake for two-week periods alternating with no treatment during the eight-week N-of-1 trial.

**Main Outcomes and Measures:** The primary outcome was discontinuation symptoms assessed twice daily by the Generic Rating Scale of Treatment Effects (0-10, with higher scores indicating greater discontinuation symptoms). Secondary outcomes were symptom expectations and depressive symptoms. Bayesian mixed models of individual and aggregated N-of-1 trial data were used to estimate posterior probabilities of treatment effect differences across different thresholds (>0 superior; ≥0.2 small effect; ≥0.8 clinically meaningful effect). Analyses were conducted as intention-to-treat.

**Results:** A total of 25 patients (mean [SD] age, 43.2 [16.0] years; 80% females) with moderate to severe discontinuation symptom scores at baseline (mean [SD], 5.8 [1.4] severity) were enrolled. Among 21 patients analyzed at individual level, 13 showed improvements in discontinuation symptoms during OLP vs no treatment, 8 indicated small and none clinically meaningful effects. Aggregated Bayesian mixed models estimated a 76% posterior probability for a superior treatment effect, with a 53% for a small and 4% for a clinically meaningful effect. Overall discontinuation symptom burden was low and decreased over time. Adverse events did not differ significantly between OLP and no treatment; there were no serious adverse events.

**Conclusion and Relevance:** This series of N-of-1 trials found OLP may improve discontinuation symptoms following clinician-supervised antidepressant discontinuation, but effects did not reach the threshold for clinical meaningfulness. Small effects in over one-third of patients indicate OLP may be a low-risk intervention for certain individuals after discontinuation.

**Trial registration:** ClinicalTrials.gov Identifier: NCT05051995 – including a priori published SAP dated December 21, 2023.

## Introduction

Discontinuation of antidepressant medication can cause adverse effects, impeding the discontinuation process. Antidepressant discontinuation symptoms are highly variable including affective and somatic symptoms.^1–3^ Meta-analyses imply that the average incidence of discontinuation symptoms ranges from 31-56%, and some individuals may experience these symptoms as severe.^1,4,5^ Symptoms typically emerge within two weeks and subside within one month, though long-term follow-ups suggest longer persistence.^5^ Evidence is lacking regarding strategies for reducing antidepressant discontinuation symptoms, yet slow tapering has been recommended.^6^

Various factors are likely to affect the risk of discontinuation symptoms, including duration of antidepressant use, maintenance dose, younger age, female sex, and symptom severity during previous discontinuation.^4,5,7–9^ Qualitative evidence suggest that earlier unsuccessful discontinuation attempts can induce fears of discontinuation symptoms and relapse.^10^ A recent meta-analysis implies that about half of antidepressant discontinuation symptoms may be attributable to nocebo effects, arising from dysfunctional expectations or non-specific symptoms,^4^ corroborated by antidepressant trials revealing strong nocebo effects.^11^ The role of expectations is further supported by meta-analytic evidence suggesting that up to 80% of the antidepressant treatment effect is also achieved within the placebo groups.^12–14^ Emergent evidence suggests that placebo effects can be induced without deception; medium to large effect sizes have been reported for open-label placebo (OLP) treatment in clinical conditions such as cancer-related fatigue, chronic pain, and major depressive disorder, among others.^15–18^

Building on these findings, we explore the effect of OLP in reducing antidepressant discontinuation symptoms. The N-of-1 trial study design is specifically suitable for studying heterogenous conditions and hard-to-reach patient groups.^19^ Single N-of-1 trials can be aggregated to generate evidence at population level, while simultaneously allowing the assessment of efficacy at individual level, thereby maximizing power for studies with relatively few participants.^20^ This study uses a series of N-of-1 trials to estimate the efficacy of OLP in patients that report discontinuation symptoms after clinician-supervised antidepressant discontinuation. We hypothesized that OLP is more efficacious than no treatment in reducing discontinuation symptoms (primary), negative symptom expectations, and depressive symptoms (secondary) during an eight-week N-of-1 trial, at both individual and population level.

## Methods

### Clinical Trial Design and Patients

This study is part of a collaborative research center (CRC; TRR 289 Treatment Expectation: treatment-expectation.de/en/) and was conducted at the University Medical Centre Hamburg-Eppendorf and the University Hospital Marburg, Germany. We performed a series of randomized, single-blind N-of-1 trials in patients (aged 18+ years) with remitted major depressive disorder after successful antidepressant discontinuation (in-and exclusion criteria provided in eTable 1 in Supplement 2). Each 8-week N-of-1 trial consisted of alternating 2-week periods OLP (A) and no treatment (B), in an ABAB or BABA sequence. The eligibility criteria, intervention protocol, and outcome measurements were identical to those used in the published study protocol.^21^ This study was approved by the ethics committee of the ‘Ärztekammer’ of Hamburg, Germany (approval number: PV7151).

### Randomization and Masking

Patients were randomly allocated to begin with either OLP or no treatment, following the ABAB or BABA scheme. Computer-generated randomization was performed via R-Studio. Assessments at study visits were conducted by blinded research assistants. Due to the nature of the study, patients, and study staff responsible for randomization were not blinded.

### Procedures Discontinuation process

Prior to the N-of-1 trial, patients completed a clinician-supervised, guideline-based^22^ antidepressant discontinuation process as part of the FAB-study,^21^ which included weekly study visits for assessments and clinical monitoring. Discontinuation began with a 1-week run-in phase at maintenance dose, followed by a 4-week tapering period using specifically manufactured capsules containing individualized doses of patients’ original medication. Discontinuation schedules consisted of 5 dose reductions, with smaller steps as doses approached zero, following hyperbolic discontinuation principles^6^ and national guidelines in effect at study initiation.^21,22^ Patients who reported at least moderate discontinuation symptoms after successful discontinuation were eligible for inclusion in the N-of-1 trial.^21^

### N-of-1 trial

Following randomization, patients entered the 8-week N-of-1 trial. The OLP period consisted of twice-daily placebo intake, with identical looking capsules as during discontinuation. A standardized verbal and written rational at the start of each OLP period aimed to convey that (1) the placebo effect can be powerful; (2) the body can automatically react to taking placebos; (3) a positive attitude helps but is not necessary; (4) taking the pills faithfully is critical.^23^ We added a fifth point: (5) an intake ritual may be helpful to foster the placebo effect. During no treatment periods, patients received no intervention to cope with discontinuation effects. Ambulatory assessment of primary and secondary outcomes was conducted twice daily with the ‘StudyU app’^24^ on the patient’s smartphone.

### Outcomes

The primary outcome was the discontinuation symptom score using a single item of the Generic Rating Scale for Treatment Effects (GEEE_ACT_),^25^ modified for ambulatory assessment. Patients rated the intensity of the experienced discontinuation symptoms during the previous 12 hours on an 11-point numeric rating scale (NRS; 0-10; higher scores indicate greater discontinuation symptoms).

Secondary outcomes included symptom expectations assessed with a modified, single item of the Generic Rating Scale for Treatment Expectations (GEEE_EXP_)^25^ (NRS; 0-10; higher scores indicate more negative symptom expectations) and depressive symptoms using the Patient-Healthcare-Questionnaire-2 (PHQ-2_SUMSCORE_)^26^ (total score 0-6; higher scores indicate more severe depressive symptoms).

Prior discontinuation experience was assessed at study start using a modified version of the Generic Rating Scale for Treatment Pre-Experiences (GEEE_PRE_),^25^ calculated as the difference between worsening and improvement ratings (NRS −10-10), with lower scores indicating more negative prior experiences. Patients without prior discontinuation were scored ‘0’. Discontinuation symptoms during tapering were measured at 4 study visits during the discontinuation process using the 43-item Discontinuation Emergent Signs and Symptoms Scale (DESS_TAPERING_)^27^ including an additional item ‘body zaps/brain zaps’.^28^ Each item was rated from 0-3; higher scores indicate more severe symptoms during tapering. Self-reported age (years), gender (female, male, non-binary), duration of antidepressant use (years), and maintenance dose (normalized to the maximum recommended dose) were collected at study start.

### Sample Size and Power

The sample size was calculated with the Shiny-App available at https://jiabeiyang.shinyapps.io/SampleSizeNof1/ which implements a linear mixed model specifically designed for N-of-1 trials.^29^ Based on this linear mixed model, a sample size of 18 patients yields a power of 93% for identifying a 0.8-point mean difference between OLP and no treatment at population level, with a significance level of 5%. Considering possible dropout, we recruited 25 patients into the study.^21^

### Statistical Analyses

The statistical analysis plan was published before data analysis on clinicaltrials.gov under the registration NCT05051995.^30^ We used Bayesian models to make inference on the treatment effect of OLP by estimating the probability of it exceeding pre-defined thresholds of the mean symptom score difference that reflect clinical meaningfulness (>0 = ‘superior’; ≥0.8 = ‘clinically meaningful’). Given that the intensive clinical support throughout our study led to milder discontinuation symptoms than anticipated, we believe that a reduction of 0.2 (‘small effect’) in the symptom score may already be clinically meaningful for patients. Hence, we also report posterior probabilities based on this threshold – defined after initial data inspection – in the statistical analysis. Intention-to-treat analyses included all randomized patients.

### Bayesian Analysis

We first analyzed the effect of OLP vs no treatment at individual level, based on the average ratings of discontinuation symptoms. A model with a linear time trend, first-order autoregressive (AR1) error structure, wide normal priors (N[0,10^-6^]) for fixed effects and uniform priors (Unif[0.1,10]) for random effects were used (eMethods 1 in Supplement 2). Separate analyses were conducted for each patient. We estimated the posterior mean treatment effect, 95% credible intervals (CrIs), and the probability of clinical meaningfulness. OLP was considered superior (and have a small/clinically meaningful effect, respectively) to no treatment if the probability of reaching the pre-defined thresholds was >50%.

Next, we examined the effect of OLP vs no treatment at population level using a Bayesian mixed model comparing the average ratings of discontinuation symptoms. In the basic model, treatment was considered as fixed effect and patients as random effect, with noninformative priors for all outcomes (eMethods 2 in Supplement 2). This model was extended with an AR1 error structure, a linear time trend, and between-patient-covariates i.e., prior experience with discontinuation, and discontinuation symptoms during tapering (extended model). Further covariates were added exploratively to assess the influence on the treatment effect (i.e., age, gender, maintenance dose, duration of use). We assumed normally distributed mean differences and estimated the posterior mean treatment effect (OLP vs no treatment), the 95% CrI, and the probability of clinical meaningfulness. Markov Chain Monte Carlo (MCMC) sampling was performed using JAGS (Just Another Gibbs Sampler) software run from R version 4.4.0. In the sampling procedure, we used 3 chains, 5000 burn-in steps, no thinning and 1000 iterations.^31^ Convergence of the models was checked using trace plots and effective sample sizes. The scripts for analysis are available at https://github.com/HIAlab/Nof1-OLP. Secondary outcomes were analyzed as detailed above for the primary outcome.

## Results

### Recruitment and Patient flow

Eligible patients were recruited from December 2021 to December 2023; with first patient-in May 11, 2022, and last-patient-out February 21, 2024. Of 62 screened patients, 28 were eligible for the N-of-1 trial, while 34 reported none or mild discontinuation symptoms following tapering and were not eligible.^21^ 3 withdrew before data collection, resulting in 25 included patients (**Figure 1**). Of these, 2 patients restarted antidepressant use, defined as major protocol violation in the statistical analysis plan^30^: data recorded until the point of antidepressant re-use were included in the analyses.

**Figure 1.**
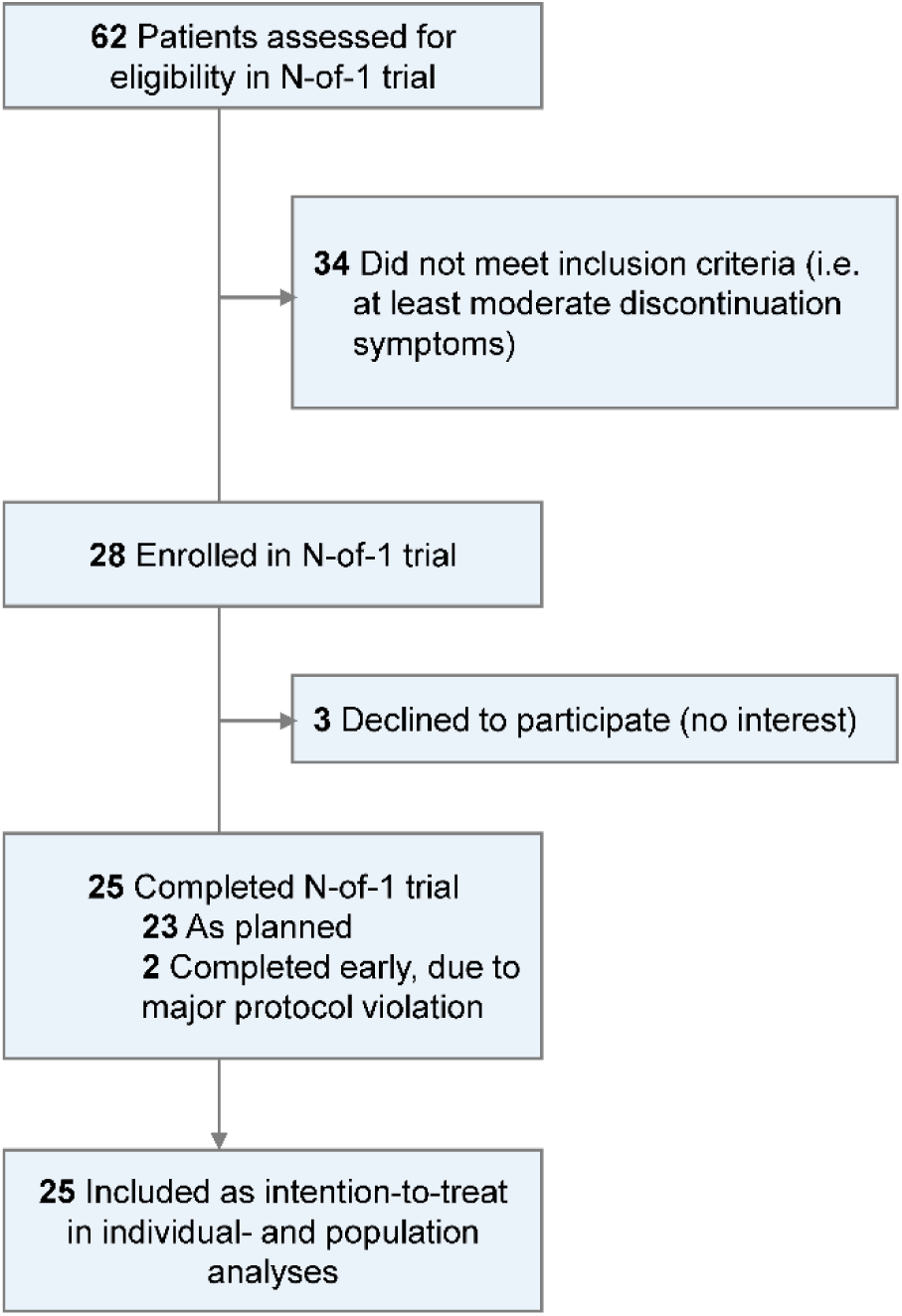
Recruitment and Patient Flow.

### Patient Characteristics

25 patients (20 females, 5 males) commenced an N-of-1 trial. **Table 1** summarizes the patient characteristics. 12 patients were randomized to ABAB and 13 to BABA. Patients completed 1795 of 2800 possible assessments (64%) of primary and secondary outcomes; 922 (51%) were collected during OLP and 873 (49%) during no treatment. The mean number of assessments per patient was 71 (possible range 1-112), with 17 (68%) showing >70% compliance (eTable 2 in Supplement 2).

**Table 1.**
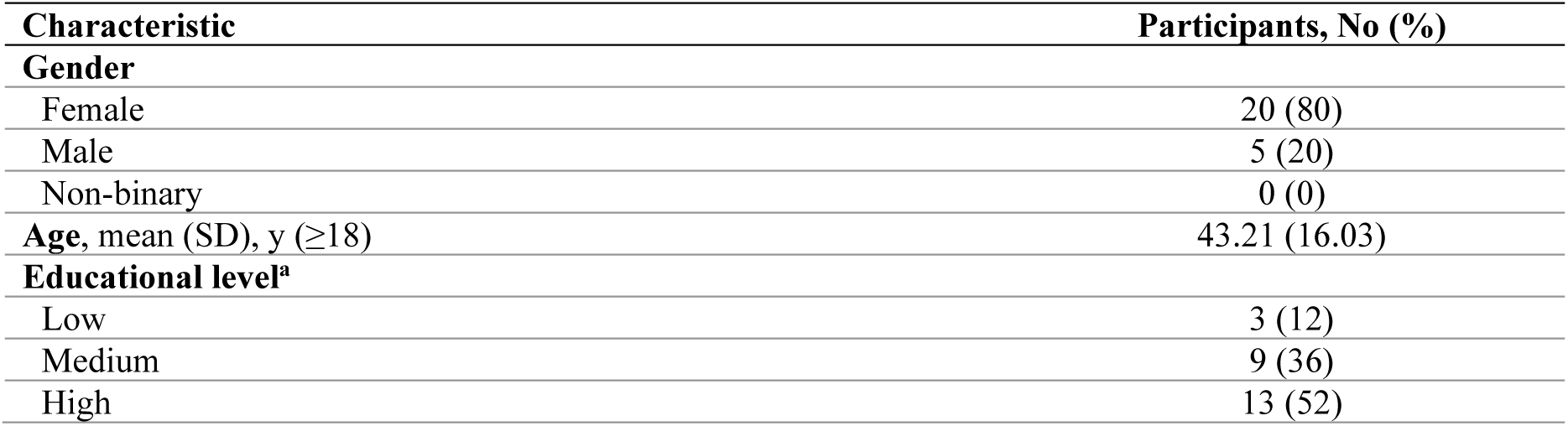
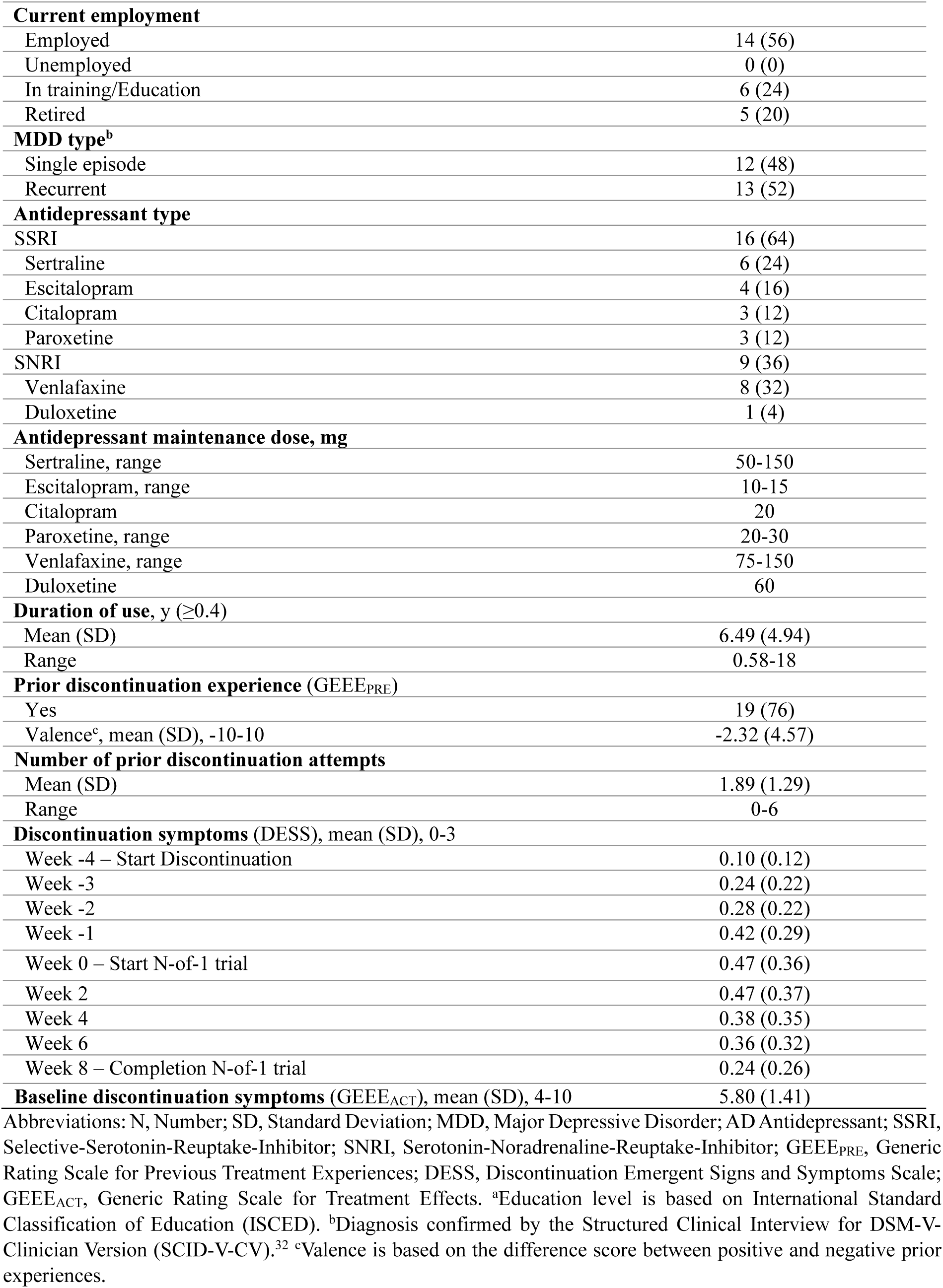
Patient Characteristics.

### Individual- and Population-Level Analyses of Primary Outcome Measure

First, we report results of the individual-level model. Quality checks showed non-converging MCMC chains for 4 patients (1, 3, 16, 22; eFigure 1 in Supplement 2) and as a result, their posterior estimates were unstable and could not be interpreted; these 4 patients were excluded from the individual-level analyses (**Figure 2**). Of the remaining 21 patients, 13 (62%) showed a >50% posterior probability of a superior OLP effect, 8 (38%) indicated a small and no patient showed a clinically meaningful effect (**Figure 3**; eTable 3 in Supplement 2).

**Figure 2.**
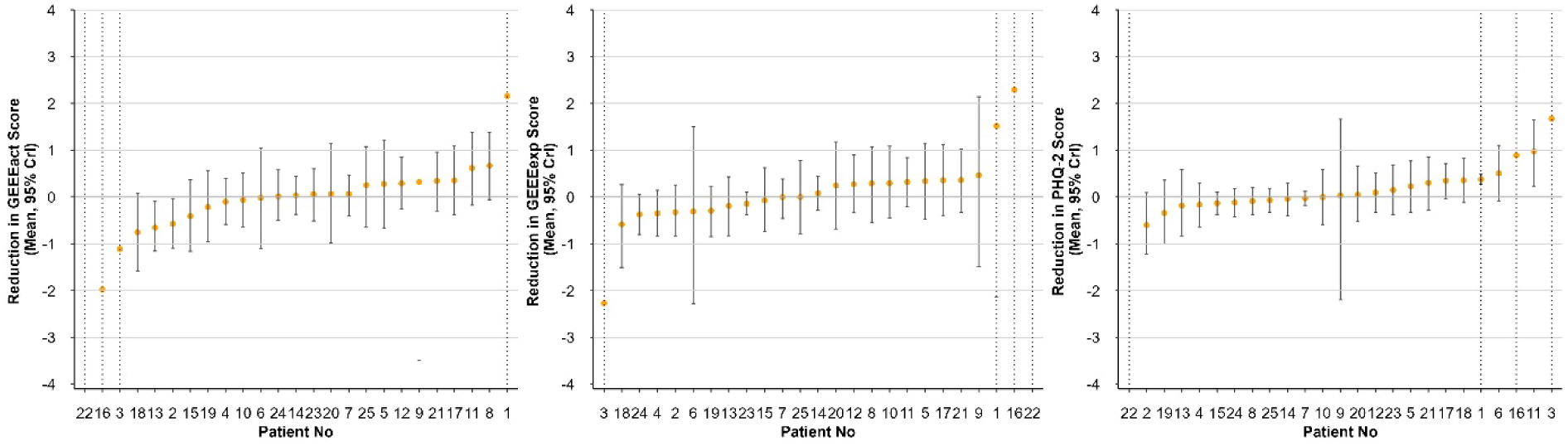
Estimated Mean Differences between Open-Label Placebo and No Treatment in Individual N-of-1 Trials. Estimated mean differences in discontinuation symptom scores (GEEE_ACT_, left), symptom expectations (GEEE_EXP_, middle) and depressive symptoms (PHQ-2, right), with 95% credible intervals, are shown for each patient (orange circles) comparing open-label placebo to no treatment. Positive values indicate greater symptom reduction with open-label placebo. Treatment effects are ordered from the smallest to the largest. Dotted lines for Patient No 1, 3, 16 and 22 (dotted lines) represent model convergence issues. Abbreviations: GEEE_ACT_, Generic Rating Scale for Treatment Effects; GEEE_EXP_, Generic Rating Scale for Treatment Expectations; PHQ-2, Patient-Healthcare-Questionnaire-2.

**Figure 3.**
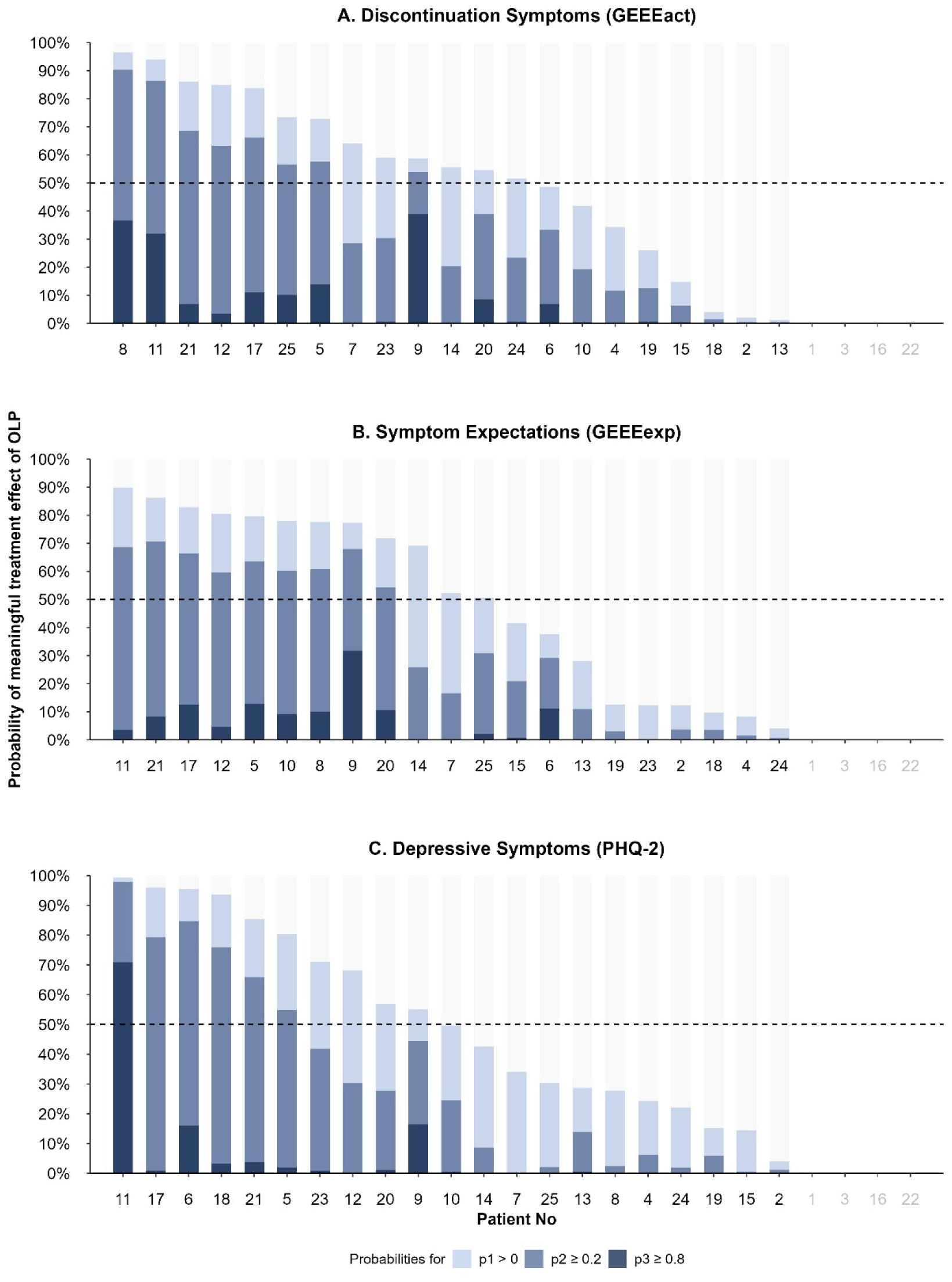
Estimated Posterior Probability of Open-Label-Placebo Treatment Effect for Individual N-of-1 Trials. Posterior probabilities of symptom improvement during OLP relative to no treatment for individual N-of-1 trials: (A) discontinuation symptoms (GEEE_ACT_), (B) symptom expectations (GEEE_EXP_), and (C) depressive symptoms (PHQ-2). Each column represents the posterior probability of a superior treatment effect of OLP compared to no treatment: p1 (light blue) representing a superior treatment effect; p2 (medium blue) a small and p3 (dark blue) a clinically meaningful effect. Effects are ordered by the probability of a superior OLP treatment effect (p1). The dotted line indicates whether this probability exceeds 50%. For four patients (1,3,16,22) the models did not converge. Abbreviations: OLP, Open-Label Placebo; GEEE_ACT_, Generic Rating Scale for Treatment Effects; GEEE_EXP_, Generic Rating Scale for Treatment Expectations; PHQ-2, Patient-Healthcare-Questionnaire-2.

**Figure 4.**
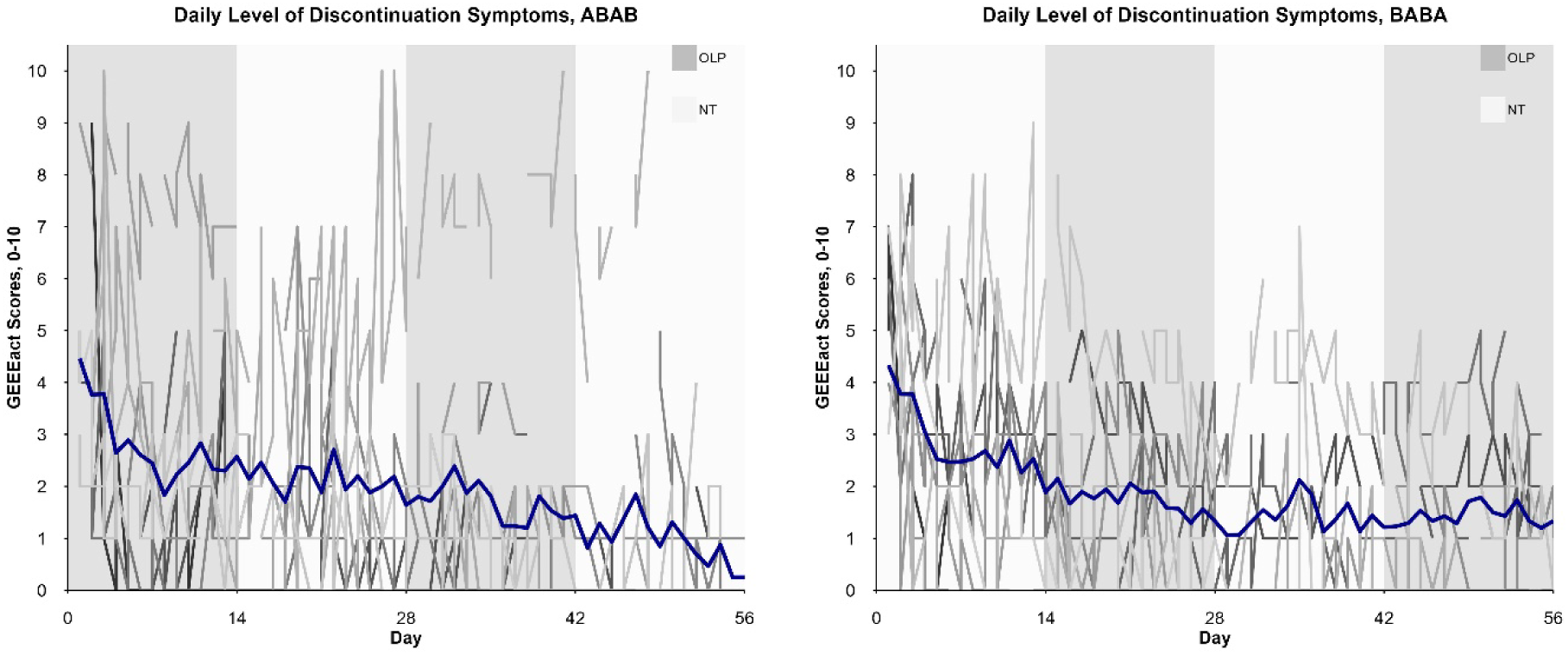
Course of Discontinuation Symptoms during Open-Label Placebo (OLP) vs No Treatment (NT) Through 8-Week N-of-1 Trial. Discontinuation symptom scores (Generic Rating Scale for Treatment Effects, GEEE_ACT_) during OLP (A) and no treatment (B) in randomization group ABAB (N = 12) and BABA (N=13). Blue line reflects the mean discontinuation symptom score for all individuals (grey lines).

At population level, the posterior mean difference of discontinuation symptoms during OLP vs no treatment was 0.23 (95% CrI: −0.75 to 1.20). In the basic model, the posterior probability for a superior OLP effect was 68%, 52% for a small and 12% for a clinically meaningful effect. Results were similar in the extended model (AR1, linear time trend, covariates) with a 76% probability of a superior, 53% for a small and 4% for a clinically meaningful treatment effect. Adding further covariates to the model did not yield substantial changes to the posterior distributions (eTable 6 in Supplement 2).

### Individual- and Population-Level Analyses of Secondary Outcome Measures

At individual level, 4 patients were excluded due to a lack of model convergence. Of the remaining 21 patients, 12 (57%) showed a superior OLP effect on symptom expectations, with 9 (43%) indicating a small and none a clinically meaningful effect (**Figure 3**; eTable 4 in Supplement 2). Similarly, for depressive symptoms, 10 patients (48%) showed a posterior probability for a superior OLP effect, with 6 (29%) showing a small and 1 (5%) a clinically meaningful effect (eTable 5 in Supplement 2). At population level, the posterior mean reduction of negative symptom expectations and depressive symptoms during OLP vs no treatment was 0.28 (95% CrI: −0.71 to 1.27) and 0.13 (95% CrI: −0.54 to 0.79), respectively. For symptom expectations, the posterior probability that OLP is superior to no treatment was 72%, with 57% for a small and 15% for a clinically meaningful effect. For depressive symptoms, the posterior probability for OLP being superior was 65%, 42% for a small and 2% for clinically meaningful effect (basic models). Incorporating further covariates or modifying the regression structure (i.e., AR1 error structure, linear time trend) did not significantly affect the posterior distributions (eTable 7, eTable 8 in Supplement 2).

### Adverse Events

No serious adverse event occurred during the study. Overall, 11 of the 25 patients (44%) reported at least one adverse event that was related to study participation. The most common adverse event was dizziness, reported by 4 patients (16%). A total of 9 adverse events were reported during OLP compared to 11 during no treatment periods, with no significant differences (eTable 9 in Supplement 2). In 2 patients, the adverse events (i.e., both reported anxiety attacks) led to the restart of antidepressant use. In case of any adverse event, patients were closely monitored according to the predefined safety protocol.^21,33^

### Adherence

Of the 25 included patients, the mean placebo adherence was 89%. Adherence was less than 72% in 5 patients, which is considered as low treatment adherence.^18^ Reasons were dropout and irregular intake.

## Discussion

This series of N-of-1 trials evaluated the efficacy of OLP compared to no treatment in reducing discontinuation symptoms among patients with remitted major depressive disorder over eight weeks following antidepressant discontinuation. Our aggregated analysis indicated that OLP may have a beneficial average effect in reducing discontinuation symptoms: there was a 76% posterior probability that OLP was more efficacious than no treatment, with a 53% probability of a small and 4% of a clinically meaningful effect. Individual-level analyses revealed that approximately two thirds of patients had some benefit of OLP, with 38% achieving a small effect, yet none a clinically meaningful effect. These findings suggest that OLP holds promise as a safe, low-effort intervention in the context of antidepressant discontinuation. Performing N-of-1 trials enabled simultaneous estimation of both individual and group-level effects, offering a personalized approach to estimate the efficacy of OLP. This is the first discontinuation study, to our knowledge, to assess discontinuation symptoms, symptom expectations and depressive symptoms in detail using ambulatory assessments. Our data suggest that discontinuation symptoms were mild and decreased substantially over eight weeks after clinician-supervised antidepressant discontinuation.

A positive effect of OLP aligns with emergent evidence supporting its efficacy across various conditions.^15^ Consistent with prior research showing modest effects of OLP in patients with depression,^34–36^ more than one-third of patients in our study experienced a small effect. However, none of these patients experienced a clinically meaningful improvement of discontinuation symptoms based on our pre-defined threshold. It is important to note that symptom burden after discontinuation was relatively mild. Given prior evidence on discontinuation symptom burden and duration in long-term users,^1^ and our selection of patients with a moderate to severe baseline symptom burden, we expected a higher symptom burden in this trial. As there were no established thresholds for clinical meaningfulness for our primary outcome, we defined a pre-specified threshold based on available evidence and clinical reasoning. However, defining such thresholds remains challenging due to the dynamic changes in the field, such as a recent meta-analysis highlighting strong nocebo effects in antidepressant discontinuation.^4^ As a consequence, we added a threshold of 0.2 and present its results. In our study, patients received clinical support throughout the discontinuation process including regular study visits and app-based tracking, which may have influenced symptoms by mitigating nocebo-related responses beyond the direct effects of OLP.^37,38^ This aligns with qualitative evidence suggesting that professional support and treatment information are experienced as central to successful discontinuation.^10^ While these findings underscore the potential benefit of intensive support throughout the discontinuation process, they also suggest a role for OLP. Particularly in naturalistic settings with limited resources, OLP holds promise as easily implementable, low-risk intervention to facilitate antidepressant discontinuation.

In secondary analyses, we observed only a small effect of OLP on symptom expectations. The induction of positive treatment expectations contributes significantly to the efficacy of OLP^15^ and may partly explain its small effects on discontinuation symptoms. The potential mediating role was beyond the scope of this study and warrants examination in future research. Additionally, we incorporated an intake ritual to foster conditioning processes, another central mechanism of OLP effects. While in our trial we focused on the efficacy of OLP after discontinuation, it has been implicated that combining OLP with conditioning throughout the process may be particularly applicable to drug discontinuation.^39,40^ As such, OLP, initially a neutral stimulus, can be paired with an active medication during a learning phase, and subsequently acts as conditioned stimulus eliciting responses typically produced by the active medication.^41,42^ Findings from a conditioned OLP trial in the context of opioid reduction found promising effects concerning a reduction of opioid consumption.^39^ Integrating the conditioning approach throughout the discontinuation process may enhance OLP efficacy and overall impact.

### Limitations

This N-of-1 trial has several limitations. First, OLP was delivered following discontinuation, when patients had already received intensive support and may have developed alternative coping strategies. In a population with higher symptom burden, the effect of OLPs may be larger. Second, while we assessed expectations regarding discontinuation symptoms, we did not assess expectations toward OLP which may have influenced its efficacy.^43^ However, the high adherence to placebo intake (89%) suggests a general acceptance of OLP. Third, the threshold of 0.2 was added post hoc after observing in the data that discontinuation symptoms were smaller than anticipated and smaller effects may be clinically meaningful. All results based on this threshold should be interpreted accordingly. Finally, the persistence of OLP effects beyond the administration period remains unclear. Existing evidence on durability is limited and primarily focuses on longer-term outcomes.^44,45^ Future research should aim to clarify whether OLPs yields lasting benefits and evaluate its temporal dynamics.

### Conclusion

This is the first study to assess the efficacy of OLP in a guided antidepressant discontinuation context using the innovative N-of-1 trial design allowing important personalized insights. Our findings suggest OLP may be beneficial for selected individuals suffering from discontinuation symptoms following antidepressant discontinuation. As a low-cost, readily available intervention with minimal risk of adverse events, OLP could support the discontinuation process. The results warrant further clinical exploration particularly in naturalistic discontinuation settings.

## Author contribution

Müller had full access to all data in the study and takes responsibility for the integrity of the data and the accuracy of the data analysis.

### Concept and design

Müller, Konigorski, Nestoriuc.

### Acquisition, analysis, or interpretation of data

Müller, Konigorski, Fadai, Warren, Koschik, Liu, Falkenberg, Kircher, Nestoriuc.

### Drafting of the manuscript

Müller

### Critical revision of the manuscript for important intellectual content

Müller, Konigorski, Fadai, Warren, Koschik, Liu, Bingel, Rief, Falkenberg, Kircher, Nestoriuc.

### Statistical analysis

Müller, Konigorski, Nestoriuc.

### Obtained funding

Nestoriuc, Kircher.

## Additional Contributions

We want to thank all participating patients for their time and effort in this study. We also like to acknowledge the following people for collecting data: Henning Romberg, Marie Schuster, Paulina Hüls, Pascal Faltermeier, Lea Hemauer, Angelika Jung, & Lukas Aldag.

## Data Availability

The study protocol and statistical analysis plan are published, and the analytic code will be shared, deidentified patient data will be available upon reasonable request.
All deidentified individual patient data, protocol, and analytic code will be available after publication without end date on request to researchers with a sound proposal.

https://designer.studyu.health

https://github.com/HIAlab/Nof1-OLP

