## Supplement 1 for "Open-Label Placebos for Antidepressant Discontinuation Symptoms: A Series of N-of-1 Trials"

Study Trial Protocol

| | |
|---|---|
| Trial ID | ClinicalTrials.gov NCT05051995<br>Ethics approval (“Ärztekammer”): PV7151 (version 3, 2021) |
| Date | 18.08.2025 |
| Sponsor | DFG, German Research Foundation): TRR 289 Treatment Expectation—Project-ID 422744262 |

Protocol Amendments

| Type of deviation | Time point | Summary of changes |
|---|---|---|
| Additional threshold for clinical meaningfulness | Statistical analyses | After we inspected the descriptive data, we noticed that the average discontinuation symptoms in our trial were milder than we anticipated (*M*=1.92, *SD*=2.00). As for a mild symptom load, a smaller difference can also be clinically meaningful, we added an additional threshold ‘>0.2’. Therefore, we also report posterior probabilities based on this threshold in the analysis. |

### Background

Antidepressant discontinuation is frequently associated accompanied by a wide range of symptoms, ranging from dizziness, nausea, insomnia, to emotional blunting and anxiety.[1] The average incidence is 56% in patients with remitted major depression, with nearly half of those cases considered severe.[1] Symptoms typically emerge within days and may persist for weeks or even months.

Placebo and nocebo effects are well-documented in antidepressant trials, highlighting the importance of expectations. Open-label placebos (OLPs) – placebos administered without deception – have shown promising results across various conditions (e.g. depression[2]; migraine[3]; chronic back pain[4]; menopausal hot flushes[5]; irritable bowel syndrome[6]; see Nestoriuc and Kleine Borgmann (2020) for an overview[7]). OLPs may offer a low-risk, cost-effective and scalable intervention for symptom management in the discontinuation phase. Implicit positive expectations associated with the pill intake can lead to beneficial treatment effects, as placebo studies indicate that placebo administration can influence both psychological and neurobiological processes.[8] This makes OLPs particularly interesting in the context of medication discontinuation, where not only pharmacological effects but also the habitual act of daily pill intake play a significant role. Scientific evidence thus suggests a potential efficacy of OLPs in reducing symptoms associated with medication discontinuation. Therefore, the FAB-study investigates the efficacy of OLP on discontinuation symptoms using a case study design, specifically a N-of-1 design.[9,10] N-of-1 trials are characterized by repeated alternating interventions (multiple cross-over trials) within a single patient, aiming to determine the individual effectiveness of these interventions.[11] Individual N-of-1 trials can then be aggregated using meta-analytic approaches to derive robust conclusions at the population level.[12,13] A key advantage of the N-of-1 design is that each patient serves as their own control, allowing for generalizable conclusions with a smaller sample size compared to randomized controlled trials.[14] The statistical power of an N-of-1 trial is further enhanced by repeated measurements during the different interventions.[13]

Applied to this study, the N-of-1 design aims to capture both the individual and population effects of OLP in reducing discontinuation symptoms (primary outcome). Further objectives include the efficacy of OLP in reducing negative symptom expectations, and depressive symptoms (secondary outcomes).

### Design & procedure

The FAB study will be conducted at two centers: the University Medical Centre Hamburg-Eppendorf, Hamburg, Germany and the University Medical Centre Marburg, Marburg, Germany. Patients who meet the inclusion and exclusion criteria and consent to daily ambulatory assessments during the observation phase will be accompanied in a case study design over a 13-week discontinuation period. For the N-of-1 design that takes place during the observation phase, we aim for a target sample size of N=20.[9,14] Since a minimum moderate burden of discontinuation symptoms is an additional inclusion criteria for this part of the study, we anticipate – based on estimates of the occurrence of discontinuation symptoms – that over-recruiting by approximately 25% (i.e. N=24-30) will allow us to reach the desired sample size for the N-of-1 design.[15,16] Recruitment for the FAB study will end once the target sample size for the N-of-1 design is achieved.

**Intervention**

The treatment factor (OLP vs no treatment) is varied within individuals in 14-day periods to assess its effect on discontinuation symptoms. A block randomization is used alternation between:

(A) OLP treatment

(B) No treatment

in sequences like ABAB or BABA (Figure 1).

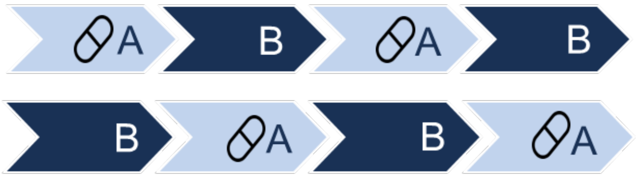

Figure 1. Schematic display of treatment sequences ABAB and BABA.

**Procedure**

The study is divided into four phases:

1. Screening and Eligibility
2. Five-week tapering (discontinuation) phase, including a one-week run-in
3. Eight-week observation phase, including the N-of-1 trial
4. Follow-up assessment, six months after baseline

(see Figure 2 for an overview)

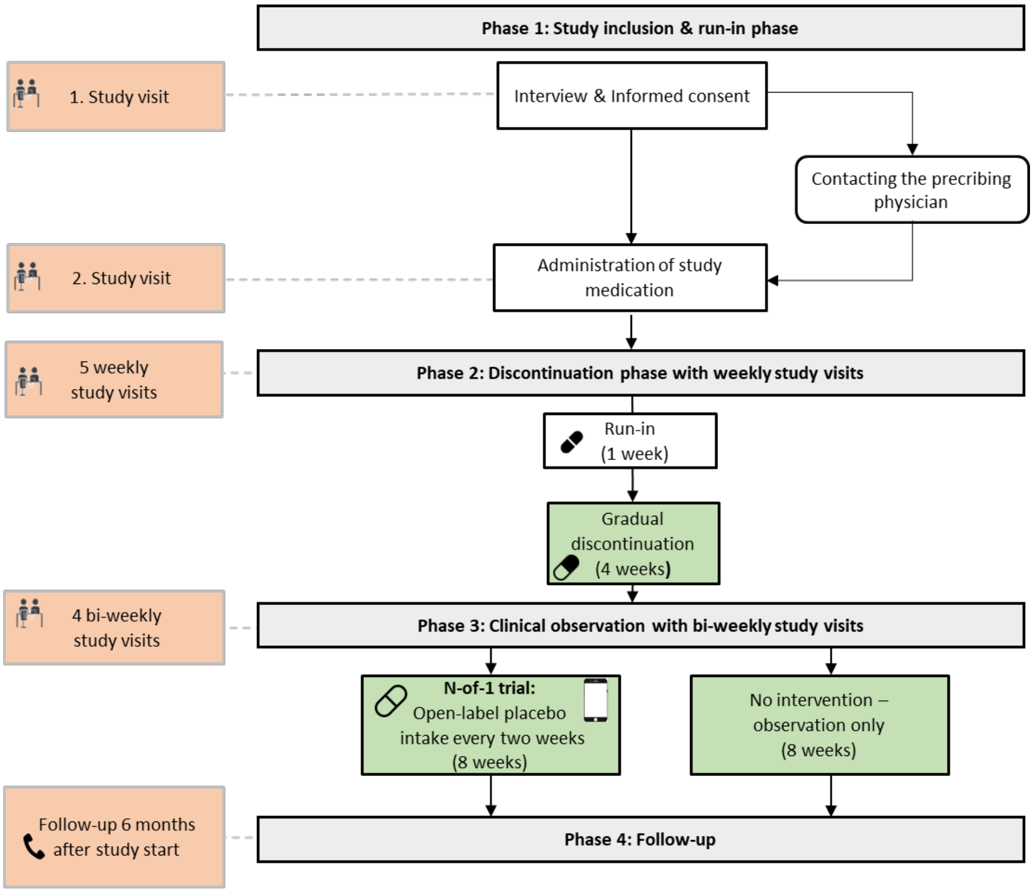

Figure 2. FAB-study procedure

##### 1. Screening & Eligibility (s1 & s2)

Individuals can access information about the study via the study website or leaflets that provided in outpatient clinics or flyer in public places (e.g. supermarket). Interested individuals can contact the research team via telephone or email. An initial telephone screening (s1; approximately 10-15 minutes) will be conducted, during which study information is provided and questions regarding sociodemographic and medical background to evaluate eligibility. Patients who meet the preliminary criteria will then be invited to an in-person screening (s2) to confirm their eligibility. This on-site screening involves patients being fully debriefed on all study procedures during the informed consent process. Subsequently, a series of clinical interviews and questionnaires to gather detailed information on sociodemographic and medical characteristics, current depressive symptoms, psychopathology and any adverse events (approximately 90 minutes) will be performed. If patients are eligible, the prescribing physician will be contacted to verify the study participation and to confirm the current medication use.

##### 2. Five-week tapering (discontinuation) phase, including a one-week run-in (t0-t5)

Following enrolment, the five-week tapering phase including a one-week run-in will begin. Patients will attend weekly study visits that include clinical interviews, online questionnaires, and the dispensing of study medication. At each study visit, safety endpoints will be systematically evaluated and monitored to ensure close supervision.

The study medication will be produced by the pharmacy at the University Medical Centre Hamburg-Eppendorf and the corresponding pharmacy at the Marburg study site, allowing for customized dose reductions. Therefore, the study medication will be packaged by trained study staff in a tabular film, with each capsule individually labelled according to the patient's tapering schedule. If a patient is unable to attend a study visit, the medication will be delivered by study staff. To accommodate for patients' availability, minor adjustments to the tapering schedule (+/- 2 days) are possible. In case of illness that interfere with the discontinuation process, the tapering period may be extended up to 2 weeks (thus a maximum of 7 weeks in total). Patients will be asked to return all unused medication to the clinic.

The tapering schedules are individually tailored based on the AD and maintenance dose of the patients. The schedule begins with a one-week run-in phase during which patients receive their antidepressant in the same dosage as their current maintenance treatment. This phase is designed to control for potential effects related to the different appearance and texture of the study medication. This is followed by a four-week tapering phase involving five dose reduction steps. The dose reductions are larger at the beginning, followed by smaller dose reductions as the dose approaches zero. This aligns with the S3 German guidelines valid at study start, recommending a four-week tapering period, and integrates evidence favoring hyperbolic dose reductions.[17] At the end of the five-week tapering schedule (t5), patients will have fully discontinued their AD, reaching a dose of zero. At this point, the eligibility for the N-of-1 trial (i.e. at least moderate discontinuation symptoms) will be assessed and patients will be informed of their group allocation by unblinded study staff. This burden is assessed using a numerical analogue scale (Generic Rating for Treatment Effects,[18] $GEEE_{ACT}$, Item 3), with a score of ≥4 indicate an at least moderate burden.

##### 3. Eight-week observation phase, including the N-of-1 trial (t6-t9)

The third phase of the study consists of the eight-week observation phase, including the N-of-1 trial for a subsample of the patients. All patients will attend bi-weekly study visits (approximately 60 minutes each), which include clinical interviews and online questionnaires. Patients who do not meet the inclusion criterion receive no intervention during the observation phase but continue to receive intensive medical and psychological support. They may also receive information about OLPs upon request.

At the start of the N-of-1 trial, eligible patients were briefed on the trial procedures by unblinded study staff. They will receive an informational leaflet outlining their treatment sequence (i.e. ABAB or BABA), detailed instructions on how to install and use the StudyU app, including information on data privacy.[19] As part of an eight-week diary assessment, they will be asked to complete short smartphone-based surveys (1-2 minutes) twice daily using the StudyU app. Each survey includes four questions addressing discontinuation symptoms, expectations, and depressive symptoms. Surveys are scheduled once in the morning (between 8.00-10.00am) and once in the evening (between 6.00-10.00pm). The StudyU app also provides information on treatment allocation (e.g. reminder for the placebo intake) and displays study progress. Push notification will remind patients to complete the assessments (two reminders per time point).

At the beginning of the first OLP period (A) a standardized OLP rationale (approximately 15 minutes) will be delivered by trained study staff based on Kaptchuk et al. 2010.[6] The rationale begins with a brief explanation of the placebo effect, including an introduction of open-label placebos, highlighting that both patients and assessors are aware that the treatment is inert. Subsequently, five discussion points will be explained:

1. The placebo effect can be powerful.
2. The body can respond automatically to placebo intake.
3. A positive attitude can help but is not required.
4. Consistent intake is crucial.
5. An intake ritual may strengthen the placebo effect.

The rationale concluded with an invitation to patients to try OLP together with the research team, as this is a novel intervention that has not yet been applied in the context of antidepressant discontinuation. Then patients will receive a leaflet summarizing these discussion points. The rationale will be briefly revisited before the second OLP phase to support understanding and adherence. At study visits patients will be provided with placebo pills that visually match the study medication and are also packages in a tabular film. The capsules will be labelled as placebo and are dispensed for a twice-daily use for a two-week period.

At the end of the eight-week observation phase (t9), patients will be informed that they finished the main part of the study and there will be only a short telephone-interview in three months. The prescribing physician will be notified about the conclusion of intensive support and will be invited to resume clinical care of the patient.

**4. Follow-up assessment (fu1), six months after baseline**

The study concludes with a follow-up six months after enrolment (t0). This assessment (approximately 45 minutes) includes clinical interviews administered online or via telephone, and online questionnaires. Patients will subsequently receive their reimbursement of up to 90 Euro.

**Participants**

Participants are patients with remitted major depression recruited from:

- The psychiatric outpatient clinic at University Medical Center Hamburg Eppendorf
- The Department of Psychiatry and Psychotherapy at Philipps-University Marburg
- General practitioners
- Flyers, leaflets, and social media

**Eligibility criteria**

*Inclusion criteria*

- Adults (18+ years) with a past diagnosis of major depressive disorder (single or recurrent), confirmed by the Structured Clinical Interview for DSM-V – Clinician Version (SCID-V-CV)[20], currently in remission
- Intake of an antidepressant with constant dosage for four weeks:
    - Citalopram 20-40mg
    - Duloxetine 60-100mg
    - Escitalopram 10-20mg
    - Paroxetine 20-40mg
    - Sertraline 50-150mg
    - Venlafaxine 75-150mg
    - Mirtazapine 30-45mg
- Wish to discontinue AD, acknowledged by the prescribing physician
- Fulfilment of the S3 guideline[21] recommendations for antidepressant discontinuation:
    - Positive response to AD
    - Full remission for ≥4 months (first episode) or ≥2 years (multiple episodes)
- Informed consent
- **Eligibility for N-of-1 trial:** Moderate to severe discontinuation symptoms after antidepressant discontinuation (t5) assessed by the $GEEE_{ACT}$[17] score ≥4/10 during past week

*Exclusion criteria*

- Current moderate/severe psychopathology
- Acute/chronic illnesses affecting depression, AD, or study participation
- Acute suicidality, psychotic symptoms, substance abuse/disorder, current mania/hypomania (SCID-V-CV)
- History of bipolar disorder or psychosis (SCID-V-CV)
- Severe stressful life events (e.g. death of a family member) within 6 months
- Insufficient German language skills

**Randomization**

Prior to study initiation, block randomization will be implemented across the two study sites to ensure balanced treatment sequences. Patients will be randomly assigned in a 1:1 ratio to one of the two treatment sequences: ABAB (OLP; no treatment; OLP; no treatment) or BABA (no treatment; OLP; no treatment; OLP). The allocation sequence will be generated using computer-based randomization in R-Studie via the blockrand package. Randomization will be performed by designated study staff who are not blinded to treatment allocation. These will also inform patients about their assigned treatment sequence and provide the corresponding instructions. Outcome assessors will remain blinded to treatment assignment throughout the study. Due to the nature of the intervention as an OLP, patients will not be blinded to their treatment allocation. They will be requested not to disclose their current treatment (OLP or no treatment) to the assessors.

**Sample size**

Sample size calculations are guided by the primary analysis assessing the efficacy of OLP compared to no treatment in reducing discontinuation symptoms, as measured by the 11-point numeric rating scale $GEEE_{ACT}$. The calculations focus on the aggregated analysis of the N-of-1 trials, which aims to estimate the average treatment effect at population level. We used the Shiny-App developed by Yang et al. (2021)[22] assessed through: https://jiabeiyang.shinyapps.io/SampleSizeNof1. This app implements a linear mixed model and is specifically designed for sample size estimation in aggregated N-of-1 trials. Our model assumed a fixed intercept and random slope model for alternating sequences (i.e., ABAB, BABA) with the following parameters:

- Number of patients: 20
- Number of assessments per patient: 112
- Residual standard deviation: 1.5
- Variance of the random slope: 0.75
- Autocorrelation (AR1 structure): 0.7

The Shiny-App supports iterative optimization to identify the minimum sample size required to meet desired statistical criteria. Based on these parameters, the results indicate that a sample size of 18 patients would provide 93% power to detect a mean difference of 0.8 points between OLP and no treatment, which we define as clinically meaningful difference, at a 5% significance level. To account for potential dropout, we plan to recruit 20 patients.

**Endpoints**

Outcomes will be assessed via clinical interviews at the study centers and via online questionnaires using Lime Survey (Lime Survey, Hamburg, Germany). During the N-of-1 trial, additional ambulatory measurements will take place via the StudyU app.[19] See Table 1 for an overview.

*Primary outcome*

*Discontinuation symptoms* will be self-reported twice daily via the StudyU app throughout the eight-week N-of-1 trial period, resulting in 112 assessments per patient. A modified, single item version of the $GEEE_{ACT}$[18] is used. For example, the evening prompt is: ‘How many side-effects caused by the discontinuation of your antidepressant medication have you experienced since this morning?’. Patients will rate symptom intensity on a 11-point numeric rating scale (NRS) from 0 (‘no side-effects’) to 10 (‘greatest side-effects imaginable’). Higher scores indicate more severe discontinuation symptoms.

*Secondary outcomes*

*Symptom expectations* will be self-reported twice daily via the StudyU app throughout the eight-week N-of-1 trial period using a modified, single item version of the $GEEE_{EXP}$.[18] Patients are asked to rate their expectations regarding discontinuation symptoms. The morning prompt is: ‘How many side-effects caused by the discontinuation of your antidepressant medication do you expect until this evening?'. These symptom expectations are rated on a 11-point NRS ranging from 0 ('no side-effects') to 10 ('greatest side-effects imaginable'). Higher scores indicate more negative symptom expectations.

*Depressive symptoms* will be self-reported twice daily via the StudyU app during the eight-week N-of-1 trial period. These assessments use the Patient-Healthcare-Questionnaire-2 (PHQ-2)[23], which comprises two items (i.e. depressed mood, anhedonia) rated on a 4-point Likert scale ranging from 0 ('not at all') to 3 ('nearly all the time'). Sum scores range from 0 to 6 with higher scores indicating greater levels of depressive symptoms.

*Effect modifier*

Prior negative experience with discontinuation and discontinuation symptom burden during tapering (t2-t5) will be assessed as possible effect modifier.

*Prior negative experiences with discontinuation* will be assessed with a modified version of the $GEEE_{PRE}$[18] at study start (t0). First, patients will be asked if they have any experience with discontinuation. Then, the assessment will consist of two items (i.e. positive/negative) concerning previous experience with antidepressant discontinuation. As an example, patients are asked: 'How positive has your experience been with discontinuing antidepressants?". Both will be rated on a 11-point NRS ranging from 0 ('no negative experience'/'no positive experience) to 10 ('maximum negative experience'/'maximum positive experience'). To calculate prior negative experience the scores of the positive experience will be subtracted from the negative experience. Patients with no prior experience will be assigned a value of 0. Sum scores range from -10 to 10 with lower scores indicating more negative prior experience with discontinuation.

*Discontinuation symptom burden during tapering* (t2-t5) will be assessed with the Discontinuation Related Signs and Symptoms Scale (DESS)[24] at four study visits during antidepressant tapering. Patients self-reported 43 discontinuation symptoms and rated the intensity of these on a 4-point scale from 0 ('not present') to 3 ('severe'). An additional item "brain/body zaps" was added.[25] Each week they will be asked to report the discontinuation symptoms of the past week. Sum scores range from 0 to 129 with higher scores indicating more severe discontinuation symptom load. Mean sum scores during tapering (t2-t5) will be used for the analysis.

Further possible effect modifier include age (years), gender (female/male/diverse), duration of AD use (years), and maintenance dose. These will be assessed at the screening on site (s2) via self-report. The maintenance dose will be additionally confirmed by the prescribing physician and will be normalized by the maximum recommended dose.

*Further assessments*
Sociodemographic characteristics, treatment expectations, state/trait anxiety and depression, stress, side-effects of antidepressant use, and mental well-being will be assessed as part of the standard CRC/TRR 289 'treatment expectation' battery.

| Name of instrument | Construct | Purpose | Timepoint |
|---|---|---|---|
| Generic Rating Scale for Treatment Effects ($GEEE_{ACT}$) – item 3 | Discontinuation symptoms | Primary outcome measure | t6-t9 (continuously) |
| Generic Rating Scale for Treatment Expectations ($GEEE_{EXP}$) – item 3 | Symptom expectations | Secondary outcome measure | t6-t9 (continuously) |
| Patient-Healthcare-Questionnaire 2 (PHQ-2) | Depressive symptoms | Secondary outcome measure | t6-t9 (continuously) |
| Generic Rating Scale for Previous Treatment Experiences ($GEEE_{PRE}$) | Negative prior experience | Potential effect modifier | t0 |
| Discontinuation Emergent Signs and Symptoms Scale | Discontinuation symptoms during tapering | Potential effect modifier | t2-t5 |
| Demographic characteristics | Age, gender | Potential effect modifier | t0 |
| Medical characteristics | Duration of antidepressant use, maintenance dose | Potential effect modifier | s2 |
| Beck Depression Inventory-II (BDI-II) | Depressive symptoms, recurrence | Safety endpoint | s2-fu1 |
| Montgomery-Åsberg Depression Scale (MADRS) | Depressive symptoms, recurrence | Safety endpoint | s2-fu1 |
| Adverse events (single question) | Adverse events | Safety endpoint | s2-fu1 |
| Generic Rating Scale for Treatment Effects ($GEEE_{ACT}$) – item 2 & 3 | Discontinuation symptoms | Safety endpoint | t1-fu1 |
| Structured Clinical Interview for DSM-V - | Psychopathology | Screening | s2 |

| | | | |
|---|---|---|---|
| Clinician Version (SCID-V-CV) | | | |
| Adherence (single question) | Adherence (open-label placebo) | Outcome measure | t6-t9 |
| **Further assessments** | | | |
| Treatment Expectation Questionnaire (TEX-Q) | Treatment expectations | Further assessment | t0-t9 |
| Generic Rating Scale for Treatment Expectations ($GEEE_{EXP}$) | Treatment expectations | Further assessment | t0-fu1 |
| State-Trait Anxiety-Depression Inventory (STADI) | State/trait anxiety/depression | Further assessment | t0 |
| Perceived Stress Scale-10 (PSS-10) | Stress | Further assessment | t0 |
| Generic Assessment of Side-Effects (GASE) | Side-effects of antidepressant | Further assessment | t0, t9 |
| Short Warwick-Edinburgh Mental Well-Being Scale (SWEMWBS) | Mental well-being | Further assessment | t0-fu1 |
| Discontinuation Emergent Signs and Symptoms Scale | Discontinuation symptoms | Further assessment | t1-fu1 |
| Generic Rating Scale for Treatment Effects ($GEEE_{ACT}$) | Current treatment (discontinuation) effects | Further assessment | t1-fu1 |
| Adherence (single question) | Adherence (antidepressant) | Further assessment | t0-t5 |
| Past Discontinuation Emergent Signs and Symptoms Scale ($DESS_{PAST}$) | Prior treatment experience | Further assessment | t0 |
| Demographic and medical characteristics | Education, employment, nationality, living situation, reproductive health, antidepressant use, previous | Further assessment | s2, t0 |

| | | | |
|---|---|---|---|
| | discontinuation experience (e.g. number of attempts), current medication use, prescriber | | |
| Blood samples | Antidepressant medication blood serum level | Further assessment | t1, t9 |

Table 1: Overview of all instruments and assessments.

**Safety measures & monitoring**

Relevant safety endpoints were depressive symptoms, recurrence, suicidality, discontinuation symptoms, and adverse events. Depressive symptoms were evaluated via self-report with the Beck Depression Inventory-II (BDI-II)[26] and expert-rating with the Montgomery-Åsberg Depression Scale (MADRS)[27] at study visits (S2-fu1). The BDI-II comprises 21 items with 4 response options (0-3). Sum scores range from 0-63, with higher scores indicating more severe depressive symptoms. The MADRS includes 10 items with 7 intensity ratings (0-6), resulting in a sum score of 0-60. Higher scores indicate more severe depressive symptoms. Recurrence is suspected if BDI-II sum scores exceed 19 or MADRS scores exceed 21 over two sequential study visits, and will be (dis-)confirmed with the SCID-V-CV section A[20] by a study psychologist or psychiatrist. Discontinuation symptoms is based on the $\text{GEEE}_{\text{ACT}}$[18] item 2 (worsening) and item 3 (side-effects). Adverse events will be measured at each study visit (S2-fu1) by a single question 'did you experience any adverse event since our last study visit?'. Adverse events will be classified according to the Common Terminology Criteria for Adverse Events: grade 1 'mild', grade 2 'moderate', grade 3 'severe', grade 4 'life threatening', and grade 5 'death'. The causality will be assigned according to the World Health Organization: 1 'certain', 2 'probable', 3 'possible', 4 'unlikely', 5 'conditional/unclassified', and 6 'unassessable/unclassifiable'.

Safety-relevant endpoints will be evaluated at each study visit by trained study staff. The pre-defined safety plan comprises 3 safety stages: stage 1 'mild', stage 2 'moderate', stage 3 'severe' and is based on Meißner et al. (2023).[28] See Table 2. Patients may withdraw at any time for any reason. Treatment will be discontinued in case of pregnancy, withdrawal of consent, or concerns due to health risks or non-compliance. If treatment is terminated prematurely, study participation options will be discussed, and patients will be encouraged to complete all assessments. If a study visit is missed due to illness, medication will be delivered. In case where illness interferes with participation, discontinuation schedules may be extended up to seven weeks, pausing dose reduction during a maximum of 2 weeks. Patients will be asked to return all unused medication, including tabular films.

| **Stages** | **Safety endpoints (cutoffs)** | **Monitoring** |
|---|---|---|
| Stage 1 'mild' | • Mild adverse event (grade 1)<br>• Discontinuation symptoms ($\text{GEEE}_{\text{ACT}}$ item 2 or item 3 ≥ 8) | 3-week monitoring procedure with additional weekly study visits as needed. |

| | | |
|---|---|---|
| | • Suicidal ideation (BDI-II item 9 = 1 or MADRS item 10 = 1 or 2<br>• Abnormal clinical impression | |
| Stage 2 'moderate' | • Moderate adverse event (grade 2)<br>• Moderate depressive symptoms (BDI-II sumscore ≥ 20 or MADRS ≥ 22)<br>• Suicidal thoughts (BDI-II item 9 = 2 or MADRS item 10 = 3 or 4)<br>• Abnormal clinical impression | 6-week intensive monitoring with additional weekly study visits as needed. |
| Stage 3 'severe' | • Severe adverse event (grade 3)<br>• Severe depressive symptoms (BDI-II sumscore ≥ 29 or MADRS ≥ 29 for two weeks)<br>• Suicidal thoughts (BDI-II item 9 = 3 or MADRS item 10 ≥ 5)<br>• Abnormal clinical impression | Study treatment terminated and immediate psychiatric treatment initiated. |

Table 2: Schematic display of the safety-plan. Note. BDI = Becks Depression Inventory; MADRS = Montgomery-Åsberg Depression Scale; $GEEE_{ACT}$ = Generic Rating Scale for Treatment Effects.

**Statistical analysis**

The aim of this study is to examine the treatment effect of OLP compared to no treatment during the eight-week N-of-1 trial. The treatment effect will concern the reduction in discontinuation symptoms (primary outcome) during OLP treatment relative to no treatment. The primary analysis will assess the treatment effect both at the individual level (i.e., how each individual responds to the treatment) and at population level (i.e., the overall treatment effect across all patients). All analyses will be conducted using the intention-to-treat (ITT) population, thus all patients who were randomized and have at least one outcome datapoint will be included in the analysis.

For the primary analysis, we used Bayesian models to derive estimates of the treatment effect of OLP relative to no treatment. Therefore, we estimated the posterior probability of it exceeding pre-defined thresholds of the mean difference between OLP and no treatment that we considered as clinically meaningful: >0 'superior' and ≥0.8 'clinically meaningful'.

*Individual level*

Bayesian models will be used to investigate the treatment effect for the primary outcome (discontinuation symptoms) at individual level. These models will compare average symptom ratings during OLP treatment periods to those during no treatment periods for each patient individually. The model included treatment (categorical: OLP, no treatment) and time(continuous) as fixed effects. To account for the longitudinal structure of the data, we incorporate a first order autoregressive (AR1) error structure, modelling the residuals such that observations closer in time were more strongly correlated than those further apart. Non-informative priors will be used for all parameters, reflecting the absence of prior information about the expected effect size of OLP treatment in this context. Posterior distributions of the average treatment effect were computed for each individual and summary statistics (e.g. mean difference between treatments, credible interval (CrI), probability of a clinically meaningful effect) were derived. Secondary outcomes such as symptom expectations and depressed mood were analysed using the same modelling strategy.

*Population level*

Bayesian mixed models, that aggregate data across all individual N-of-1 trials, will be applied to assess the treatment effect for the primary outcome (discontinuation symptoms) at population level. These models will compare the average symptom ratings during OLP treatment periods to no treatment periods across all individuals. The modelling process begins with a basic hierarchical structure, including a fixed effect for treatment, and random intercepts for individuals. This model is then extended to incorporate a linear time trend, an AR-1 error structure, and the inclusion of between-patient covariates such as (1) prior negative discontinuation experiences, discontinuation symptoms over time, and (2) age, female gender, higher maintenance dose, duration of AD use. These covariates will be evaluated to determine potential effect modifying effects on treatment outcomes. As with the individual-level models, non-informative priors will be used for all parameters. From this aggregated Bayesian analysis, we will derive the posterior distribution of the average treatment effect and summary statistics (e.g. mean difference between treatments, CrI, probability of a clinically meaningful effect). Secondary outcomes will be analyzed using the same modelling strategy.

All statistical analyses will be conducted in JAGS, interfaced through R, using Markov Chain Monte Carlo (MCMC) methods to generate empirical samples from the joint posterior distribution of model parameters.

**Data management**

The research team plans to develop a manualized Standard Operating Procedure (SOP) to ensure consistency in clinical interviews and assessments, and to minimize errors and data loss. All data collection and handling procedures will comply with the European General Data Protection Regulation (GDPR). Patient data will be saved in pseudonymous form, which will be performed with the ALLIAS software including a two-factor authentication and a deterministic pseudonymization technique.[29] Trial-specific documents will be stored securely, with access restricted to nominated research staff. Data from digital questionnaires and smartphone-based assessments will be stored securely:

- LimeSurvey data will be stored on a secure sever provided by the University of Duisburg-Essen, with restricted cloud access for authorized staff.

- StudyU app[19] data will be encoded using individual invite codes that are not linked to any personal identifiers. Anonymized data will be stored on a secure backend hosted by the Hasso-Plattner-Institute, Potsdam, Germany, and locally at each site.

For the StudyU app patients will be asked to agree to its terms of use, which include information about data storage and publication. Patients will be informed that the app can be deleted after study completion.

*Data Safety Monitoring Board (DSMB)*
An independent DSMB will oversee the conduct of the study and advise on patient safety. The DSMB will receive bi-annual safety reports summarizing adverse events, enrolment progress, and any relevant clinical observations (safety endpoints, i.e., depressive symptoms, suicidality, recurrence, and discontinuation symptoms). Serious adverse events (SAEs), including severe depressive symptomatology, acute suicidality, or other medically significant events, will be reported to the DSMB within 48h of occurrence. The DSMB will review these reports to assess patient safety and data integrity, and will make recommendations regarding the continuation, modification, or termination of the study. All submitted data will be pseudonymized to ensure patients confidentiality.

### Implications

This trial will be the first to assess the efficacy of OLP in reducing antidepressant discontinuation symptoms using a series of N-of-1 trials. By applying Bayesian analyses, the study will estimate treatment effects both at the individual and at population level, offering in-depth insights into the effect of OLP in the context of antidepressant discontinuation. Our study design allows for robust conclusions with a smaller sample size than traditional randomized controlled trials, making it specifically feasible for hard-to-recruit populations. Standardized discontinuation schedules and comprehensive ambulatory assessments will enable more precise understanding of the temporal dynamics of discontinuation symptoms, symptom expectations, and depressive symptoms. Finally, our trial will provide evidence whether OLP can act as supportive intervention in clinical practice to facilitate antidepressant discontinuation and improve patient outcomes.
