## Supplement 2 for "Open-Label Placebos for Antidepressant Discontinuation Symptoms: A Series of N-of-1 Trials"

**Submission title and authors:**

### Content

eTable 1. Inclusion and Exclusion Criteria 2
eTable 2. Compliance to Ambulatory Assessment (Primary and Secondary Outcomes) 3
eTable 3. Results Bayesian Analysis at Individual Level for the Primary Outcome – Discontinuation Symptoms ($GEEE_{ACT}$) 4
eTable 4. Results Bayesian Analysis at Individual Level for Secondary Outcome – Symptom Expectations ($GEEE_{EXP}$) 5
eTable 5. Results Bayesian Analysis at Individual Level for Secondary Outcome – Depressive Symptoms (PHQ-2) 6
eTable 6. Results of Bayesian Analysis at Population Level for the Primary Outcome - Discontinuation Symptoms ($GEEE_{ACT}$) 7
Exploratory: 7
eTable 7. Results of Bayesian Analysis at Population Level for the Secondary Outcome Symptom Expectations ($GEEE_{EXP}$) 8
Exploratory: 8
eTable 8. Results of Bayesian Analysis at Population Level for the Secondary Outcome Depressive Symptoms (PHQ-2) 9
Exploratory: 9
eTable 9. Adverse events 11
eFigure 1. Quality Control Bayesian Analyses – Convergence of the MCMC chain 12
eMethods 1. Bayesian Model to Estimate the Treatment Effect at Individual Level 13
eMethods 2. Bayesian Mixed Model to Estimate the Treatment Effect at Population Level (Basic Model) 14

eTable 1. Inclusion and Exclusion Criteria

| **Inclusion criteria** |
|---|
| 1. Adult patients (≥18) with fully remitted MDD, single or recurrent, confirmed by the Structured Clinical Interview for DSM-V – Clinical Version (SCID-V-CV)[1] |
| 2. Antidepressant use (citalopram (20-40 mg), duloxetine (60-100 mg), escitalopram (10-20 mg), paroxetine (20-40 mg), sertraline (50-150 mg), venlafaxine (75-150 mg) or mirtazapine (30-45 mg)) with a constant dosage for four weeks |
| 3. Discontinuation wish by participant, acknowledged by prescribing physician |
| 4. Fulfilment of the S3 German national guideline recommendations to discontinue antidepressant medication: (a) response to antidepressant medication; (b) symptom remission ≥ 4 months (first episode) or ≥ 2 years (≥ 2 episodes with significant functional impairment)[2] |
| 5. Informed consent |
| 6. At least moderate discontinuation symptoms following antidepressant discontinuation assessed by the Generic Rating Scale for Treatment Effects ($GEEE_{ACT}$ score ≥ 4/10 during past week)[3] |
| **Exclusion criteria** |
| 1. Current moderate or severe psychopathological symptoms or psychosocial impairments |
| 2. Acute or chronic somatic illness which might interfere with depressive disorder, antidepressant use or proposed study |
| 3. Acute suicidality, psychotic symptoms, substance abuse or addiction, current mania or hypomania confirmed by SCID-V-CV or other psychopathology which might interfere with depressive disorder, antidepressant use or proposed study |
| 4. Any history of bipolar disorder or psychosis confirmed by SCID-V-CV |
| 5. Severe stressful life events within six months prior to study participation |
| 6. Current pregnancy |
| 7. Insufficient German language proficiency |

*Note.* The inclusion and exclusion criteria were identical to those published in the study protocol.[4]

eTable 2. Compliance to Ambulatory Assessment (Primary and Secondary Outcomes)

| Patient No | Actual observations (N) | Possible observations (N) | Compliance (%) |
|---|---|---|---|
| **25** | 106 | 0-112 | 95 |
| **23** | 105 | 0-112 | 94 |
| **24** | 105 | 0-112 | 94 |
| **14** | 104 | 0-112 | 93 |
| **7** | 103 | 0-112 | 92 |
| **15** | 101 | 0-112 | 90 |
| **12** | 99 | 0-112 | 88 |
| **2** | 98 | 0-112 | 88 |
| **5** | 98 | 0-112 | 88 |
| **17** | 98 | 0-112 | 88 |
| **21** | 98 | 0-112 | 88 |
| **8** | 90 | 0-112 | 80 |
| **18** | 83 | 0-112 | 74 |
| **10** | 82 | 0-112 | 73 |
| **11** | 82 | 0-112 | 73 |
| **4** | 79 | 0-112 | 71 |
| **13** | 77 | 0-112 | 69 |
| **19** | 61 | 0-112 | 54 |
| **6** | 35 | 0-112 | 31 |
| **20** | 35 | 0-112 | 31 |
| **3** | 23 | 0-112 | 21 |
| **9** | 17 | 0-112 | 15 |
| **1** | 12 | 0-112 | 11 |
| **16** | 3 | 0-112 | 3 |
| **22** | 1 | 0-112 | 1 |

*Note.* N = Number.

eTable 3. Results Bayesian Analysis at Individual Level for the Primary Outcome – Discontinuation Symptoms ($GEEE_{ACT}$)

| **Patient No** | **Mean treatment effect*** | **SD** | **95% Credible Interval** | **Posterior probability of a difference >0** | **Posterior probability of a difference ≥0.2** | **Posterior probability of a difference ≥0.8** |
|---|---|---|---|---|---|---|
| **8** | 0.67 | 0.37 | -0.06-1.38 | 97% | 90% | 37% |
| **11** | 0.62 | 0.40 | -0.17-1.38 | 94% | 86% | 32% |
| **21** | 0.34 | 0.32 | -0.30-0.96 | 86% | 69% | 7% |
| **12** | 0.29 | 0.28 | -0.26-0.85 | 85% | 63% | 3% |
| **17** | 0.36 | 0.37 | -0.38-1.09 | 84% | 66% | 11% |
| 1 | 2.16 | 3.56 | -6.11-9.15 | 80% | 78% | 72% |
| **25** | 0.26 | 0.43 | -0.65-1.08 | 73% | 57% | 10% |
| **5** | 0.28 | 0.48 | -0.66-1.22 | 73% | 58% | 14% |
| **7** | 0.07 | 0.22 | -0.41-0.48 | 64% | 29% | 0% |
| **23** | 0.06 | 0.28 | -0.51-0.60 | 59% | 30% | 0% |
| **9** | 0.32 | 1.89 | -3.48-4.10 | 59% | 54% | 39% |
| **14** | 0.03 | 0.20 | -0.37-0.45 | 55% | 20% | 0% |
| **20** | 0.06 | 0.54 | -0.98-1.14 | 55% | 39% | 8% |
| **24** | 0.02 | 0.27 | -0.50-0.58 | 52% | 23% | 1% |
| 16 | -1.98 | 22.59 | -54.32-42.95 | 51% | 50% | 49% |
| **6** | -0.02 | 0.55 | -1.09-1.04 | 49% | 33% | 7% |
| 3 | -1.11 | 15.29 | -30.08-28.07 | 48% | 48% | 47% |
| **10** | -0.06 | 0.30 | -0.64-0.51 | 42% | 19% | 0% |
| 22 | -4.35 | 29.02 | -60.79-53.31 | 39% | 39% | 38% |
| **4** | -0.10 | 0.25 | -0.58-0.39 | 34% | 12% | 0% |
| **19** | -0.22 | 0.37 | -0.95-0.56 | 26% | 12% | 0% |
| **15** | -0.41 | 0.39 | -1.16-0.38 | 15% | 6% | 0% |
| **18** | -0.75 | 0.43 | -1.57-0.09 | 4% | 1% | 0% |
| **2** | -0.57 | 0.27 | -1.09- -0.03 | 2% | 0% | 0% |
| **13** | -0.65 | 0.27 | -1.16- -0.09 | 1% | 0% | 0% |

*Note.* *Mean treatment effect of open-label placebo relative to no treatment in reducing discontinuation symptoms during eight-week N-of-1 trial. Effects are ordered by posterior probability of a superior treatment effect (>0). Four patients were excluded due to a lack of convergence (gray).

eTable 4. Results Bayesian Analysis at Individual Level for Secondary Outcome – Symptom Expectations ($\text{GEEE}_{\text{EXP}}$)

| **Patient No** | **Mean treatment effect*** | **SD** | **95% Credible Interval** | **Posterior probability of a difference >0** | **Posterior probability of a difference ≥0.2** | **Posterior probability of a difference ≥0.8** |
|---|---|---|---|---|---|---|
| **11** | 0.32 | 0.26 | -0.21 - 0.84 | 90% | 68% | 3% |
| **21** | 0.36 | 0.33 | -0.33 - 1.03 | 86% | 70% | 8% |
| 1 | 1.52 | 1.74 | -2.14 - 5.07 | 85% | 82% | 70% |
| **17** | 0.36 | 0.38 | -0.39 - 1.11 | 83% | 66% | 12% |
| **12** | 0.27 | 0.32 | -0.34 - 0.89 | 81% | 60% | 5% |
| **5** | 0.34 | 0.41 | -0.48 - 1.14 | 80% | 64% | 13% |
| **10** | 0.30 | 0.38 | -0.44 - 1.09 | 78% | 60% | 9% |
| **8** | 0.29 | 0.41 | -0.54 - 1.07 | 78% | 61% | 10% |
| **9** | 0.47 | 0.94 | -1.49 - 2.14 | 77% | 68% | 32% |
| **20** | 0.25 | 0.46 | -0.68 - 1.18 | 72% | 54% | 10% |
| **14** | 0.08 | 0.18 | -0.29 - 0.44 | 69% | 26% | 0% |
| 22 | 8.47 | 23.98 | -32.87 - 57.62 | 61% | 60% | 60% |
| **7** | 0.00 | 0.21 | -0.46 - 0.38 | 52% | 17% | 0% |
| 16 | 2.29 | 23.99 | -41.26 - 50.46 | 52% | 51% | 50% |
| **25** | 0.00 | 0.40 | -0.78 - 0.77 | 51% | 31% | 2% |
| 3 | -2.27 | 13.85 | -30.67 - 25.84 | 43% | 43% | 41% |
| **15** | -0.07 | 0.34 | -0.74 - 0.62 | 42% | 21% | 1% |
| **6** | -0.31 | 0.95 | -2.28 - 1.50 | 38% | 29% | 11% |
| **13** | -0.19 | 0.32 | -0.83 - 0.43 | 28% | 11% | 0% |
| **19** | -0.29 | 0.27 | -0.85 - 0.22 | 13% | 3% | 0% |
| **23** | -0.14 | 0.12 | -0.38 - 0.10 | 12% | 0% | 0% |
| **2** | -0.32 | 0.28 | -0.84 - 0.26 | 12% | 4% | 0% |
| **18** | -0.58 | 0.45 | -1.50 - 0.26 | 10% | 4% | 0% |
| **4** | -0.35 | 0.25 | -0.83 - 0.14 | 8% | 2% | 0% |
| **24** | -0.37 | 0.22 | -0.80 - 0.05 | 4% | 1% | 0% |

*Note.* *Mean treatment effect of open-label placebo relative to no treatment in reducing dysfunctional symptom expectations during eight-week N-of-1 trial. Effects are ordered by posterior probability of a superior treatment effect (>0). Four patients were excluded due to a lack of convergence (gray).

eTable 5. Results Bayesian Analysis at Individual Level for Secondary Outcome – Depressive Symptoms (PHQ-2)

| **Patient No** | **Mean treatment effect*** | **SD** | **95% Credible Interval** | **Posterior probability of a difference >0** | **Posterior probability of a difference ≥0.2** | **Posterior probability of a difference ≥0.8** |
|---|---|---|---|---|---|---|
| **1** | 0.38 | 0.05 | 0.28 - 0.49 | 100% | 100% | 0% |
| **11** | 0.98 | 0.35 | 0.23 - 1.65 | 99% | 98% | 71% |
| **17** | 0.35 | 0.19 | -0.04 - 0.72 | 96% | 79% | 1% |
| **6** | 0.51 | 0.30 | -0.09 - 1.10 | 96% | 85% | 16% |
| **18** | 0.36 | 0.24 | -0.12 - 0.82 | 94% | 76% | 3% |
| **21** | 0.30 | 0.29 | -0.28 - 0.85 | 85% | 66% | 4% |
| **5** | 0.23 | 0.28 | -0.32 - 0.77 | 80% | 55% | 2% |
| **23** | 0.15 | 0.27 | -0.38 - 0.68 | 71% | 42% | 1% |
| **12** | 0.10 | 0.21 | -0.33 - 0.52 | 68% | 30% | 0% |
| **3** | 1.68 | 13.27 | -26.50 - 25.27 | 59% | 58% | 57% |
| **20** | 0.05 | 0.30 | -0.52 - 0.66 | 57% | 28% | 1% |
| **9** | 0.03 | 0.97 | -2.20 - 1.66 | 55% | 45% | 16% |
| **10** | 0.00 | 0.30 | -0.59 - 0.58 | 50% | 24% | 1% |
| **16** | 0.89 | 24.08 | -47.31 - 47.39 | 49% | 49% | 48% |
| **14** | -0.04 | 0.18 | -0.40 - 0.30 | 43% | 9% | 0% |
| **7** | -0.03 | 0.07 | -0.18 - 0.12 | 34% | 0% | 0% |
| **25** | -0.07 | 0.13 | -0.32 - 0.19 | 30% | 2% | 0% |
| **13** | -0.18 | 0.35 | -0.83 - 0.59 | 29% | 14% | 1% |
| **8** | -0.09 | 0.15 | -0.38 - 0.20 | 28% | 2% | 0% |
| **22** | -15.00 | 24.41 | -66.97 - 33.51 | 27% | 27% | 26% |
| **4** | -0.16 | 0.24 | -0.65 - 0.30 | 24% | 6% | 0% |
| **24** | -0.12 | 0.15 | -0.42 - 0.18 | 22% | 2% | 0% |
| **19** | -0.34 | 0.34 | -1.00 - 0.36 | 15% | 6% | 0% |
| **15** | -0.13 | 0.13 | -0.37 - 0.11 | 15% | 1% | 0% |
| **2** | -0.60 | 0.33 | -1.22 - 0.09 | 4% | 1% | 0% |

*Note.* *Mean treatment effect of open-label placebo relative to no treatment in reducing depressive symptoms during eight-week N-of-1 trial. Effects are ordered by posterior probability of a superior treatment effect (>0). Four patients were excluded due to a lack of convergence (gray).

eTable 6. Results of Bayesian Analysis at Population Level for the Primary Outcome - Discontinuation Symptoms (GEEE$_{ACT}$)

**Primary model:**

Fixed: Intervention, Time, Prior Discontinuation Experience (GEEE$_{PRE}$), Discontinuation Symptoms During Tapering (DESS$_{TAPERING}$)

Random: Patients

Autocorrelation: AR1

| **Parameter** | **Mean** | **SD** | **CrI (95%)** |
|---|---|---|---|
| Beta DESS$_{TAPERING}$ | 3.57 | 0.94 | 1.66 - 5.36 |
| Beta GEEE$_{PRE}$ | -0.15 | 0.05 | -0.24 - -0.06 |
| Beta Time | -0.03 | 0.01 | -0.05 - 0.00 |
| Difference (NT-OLP) | 0.23 | 0.33 | -0.41 - 0.89 |
| MU NT | 1.21 | 0.42 | 0.39 - 2.04 |
| MU OLP | 0.98 | 0.41 | 0.19 - 1.82 |
| P1 (>0) | 0.76 | 0.43 | 0.00 - 1.00 |
| P2 (≥0.2) | 0.53 | 0.50 | 0.00 - 1.00 |
| P3 (≥0.8) | 0.04 | 0.20 | 0.00 - 1.00 |
| Phi | 0.53 | 0.03 | 0.48 - 0.58 |
| Sigma NT | 1.01 | 0.23 | 0.65 - 1.54 |
| Sigma OLP | 0.92 | 0.20 | 0.59 - 1.40 |
| Sigma within | 1.23 | 0.02 | 1.19 - 1.28 |

Abbreviations: GEEE$_{PRE}$ = Generic Rating Scale for Treatment Effects (previous treatment experiences); DESS$_{TAPERING}$ = Discontinuation Emergent Signs and Symptoms Scale (during antidepressant tapering); AR1 = First-Order Autoregressive Error Structure; NT = No Treatment; OLP = Open-Label Placebo; P1 = Posterior Probability of Treatment Effect >0; P2 = Posterior Probability of Treatment Effect ≥0.2; P3 = Posterior Probability of Treatment Effect ≥0.8; CrI = Credible Interval.

Exploratory:

Fixed: Intervention, Time, Prior Experience (GEEE$_{PRE}$), Discontinuation Symptoms During Tapering (DESS$_{TAPERING}$), Maintenance Dose, Duration of Use, Age, Gender

Random: Patients

Autocorrelation: AR1

| **Parameter** | **Mean** | **SD** | **CrI** |
|---|---|---|---|
| Beta Maintenance Dose | -0.12 | 0.36 | -0.82 - 0.59 |
| Beta Duration of Use | -0.09 | 0.03 | -0.16 - -0.02 |
| Beta Age | 0.01 | 0.01 | -0.01 - 0.03 |
| Beta Gender | 1.26 | 0.44 | 0.36 - 2.12 |
| Beta DESS$_{TAPERING}$ | 4.83 | 0.93 | 2.97 - 6.63 |
| Beta GEEE$_{PRE}$ | -0.16 | 0.05 | -0.26 - -0.07 |
| Beta Time | -0.03 | 0.01 | -0.05 - -0.01 |
| Difference (NT-OLP) | 0.22 | 0.30 | -0.36 - 0.80 |
| MU NT | 0.05 | 0.76 | -1.39 - 1.65 |
| MU OLP | -0.17 | 0.75 | -1.59 - 1.39 |
| P1 (>0) | 0.77 | 0.42 | 0.00 - 1.00 |
| P2 (≥0.2) | 0.52 | 0.50 | 0.00 - 1.00 |
| P3 (≥0.8) | 0.03 | 0.16 | 0.00 - 1.00 |
| Phi | 0.53 | 0.03 | 0.48 - 0.58 |
| Sigma NT | 0.91 | 0.22 | 0.56 - 1.41 |
| Sigma OLP | 0.77 | 0.19 | 0.47 - 1.20 |
| Sigma within | 1.23 | 0.02 | 1.19 - 1.28 |

Abbreviations: GEEE$_{PRE}$ = Generic Rating Scale for Treatment Effects (previous treatment experiences); DESS$_{TAPERING}$ = Discontinuation Emergent Signs and Symptoms Scale (during antidepressant tapering); AR1 = First-Order Autoregressive Error Structure; NT = No Treatment; OLP

= Open-Label Placebo; P1 = Probability of Treatment Effect >0; P2 = Probability of Treatment Effect ≥0.2; P3 = Posterior Probability of Treatment Effect ≥0.8; CrI = Credible Interval.

eTable 7. Results of Bayesian Analysis at Population Level for the Secondary Outcome Symptom Expectations (GEEE$_{EXP}$)

**Primary model:**

Fixed: Intervention, Time, Prior Experience (GEEE$_{PRE}$), Discontinuation Symptoms During Tapering (DESS$_{TAPERING}$)

Random: Patients

Autocorrelation: AR1

| **Parameter** | **Mean** | **SD** | **CrI** |
|---|---|---|---|
| Beta DESS$_{TAPERING}$ | 3.57 | 0.94 | 1.66 - 5.36 |
| Beta GEEE$_{PRE}$ | -0.15 | 0.05 | -0.24 - -0.06 |
| Beta Time | -0.03 | 0.01 | -0.05 - 0.00 |
| Difference (NT-OLP) | 0.23 | 0.33 | -0.41 - 0.89 |
| MU NT | 1.21 | 0.42 | 0.39 - 2.04 |
| MU OLP | 0.98 | 0.41 | 0.19 - 1.82 |
| P1 (>0) | 0.76 | 0.43 | 0.00 - 1.00 |
| P2 (≥0.2) | 0.53 | 0.50 | 0.00 - 1.00 |
| P3 (≥0.8) | 0.04 | 0.20 | 0.00 - 1.00 |
| Phi | 0.53 | 0.03 | 0.48 - 0.58 |
| Sigma NT | 1.01 | 0.23 | 0.65 - 1.54 |
| Sigma OLP | 0.92 | 0.20 | 0.59 - 1.40 |
| Sigma within | 1.23 | 0.02 | 1.19 - 1.28 |

Abbreviations: GEEE$_{PRE}$ = Generic Rating Scale for Treatment Effects (previous treatment experiences); DESS$_{TAPERING}$ = Discontinuation Emergent Signs and Symptoms Scale (during antidepressant tapering); AR1 = First-Order Autoregressive Error Structure; NT = No Treatment; OLP = Open-Label Placebo; P1 = Posterior Probability of Treatment Effect >0; P2 = Posterior Probability of Treatment Effect ≥0.2; P3 = Posterior Probability of Treatment Effect ≥0.8; CrI = Credible Interval.

Exploratory:

Fixed: Intervention, Time, Prior Experience (GEEE$_{PRE}$), Discontinuation Symptoms During Tapering (DESS$_{TAPERING}$), Maintenance Dose, Duration of Use, Gender

Random: Patients

Autocorrelation: AR1

| **Parameter** | **Mean** | **SD** | **CrI** |
|---|---|---|---|
| Beta Maintenance Dose | -0.23 | 0.37 | -0.96 - 0.51 |
| Beta Duration of Use | -0.11 | 0.04 | -0.18 - -0.04 |
| Beta Age | 0.01 | 0.01 | -0.02 - 0.03 |
| Beta Gender | 1.43 | 0.49 | 0.48 - 2.41 |
| Beta DESS$_{TAPERING}$ | 5.64 | 0.97 | 3.73 - 7.58 |
| Beta GEEE$_{PRE}$ | -0.12 | 0.05 | -0.22 - -0.02 |
| Beta Time | -0.03 | 0.01 | -0.05 - -0.01 |
| Difference (NT-OLP) | 0.29 | 0.31 | -0.33 - 0.91 |
| MU NT | -0.02 | 0.81 | -1.66 - 1.57 |
| MU OLP | -0.31 | 0.81 | -1.93 - 1.31 |
| P1 (>0) | 0.82 | 0.38 | 0.00 - 1.00 |
| P2 (≥0.2) | 0.61 | 0.49 | 0.00 - 1.00 |
| P3 (≥0.8) | 0.05 | 0.22 | 0.00 - 1.00 |
| Phi | 0.64 | 0.02 | 0.60 - 0.69 |
| Sigma NT | 0.91 | 0.23 | 0.52 - 1.44 |
| Sigma OLP | 0.85 | 0.21 | 0.51 - 1.32 |
| Sigma within | 1.05 | 0.02 | 1.01 - 1.09 |

Abbreviations: GEEE$_{PRE}$ = Generic Rating Scale for Treatment Effects (previous treatment experiences); DESS$_{TAPERING}$ = Discontinuation Emergent Signs and Symptoms Scale (during antidepressant tapering); AR1 = First-Order Autoregressive Error Structure; NT = No Treatment; OLP = Open-Label Placebo; P1 = Posterior Probability of Treatment Effect >0; P2 = Posterior Probability of Treatment Effect ≥0.2; P3 = Posterior Probability of Treatment Effect ≥0.8; CrI = Credible Interval.

eTable 8. Results of Bayesian Analysis at Population Level for the Secondary Outcome Depressive Symptoms (PHQ-2)

**Primary model:**

Fixed: Intervention, Time, Prior Experience (GEEE$_{PRE}$), Discontinuation Symptoms During Tapering (DESS$_{TAPERING}$)

Random: Patients

Autocorrelation: AR1

| Parameter | Mean | SD | CrI |
|---|---|---|---|
| Beta DESS$_{TAPERING}$ | 3.10 | 0.73 | 1.64 - 4.52 |
| Beta GEEE$_{PRE}$ | -0.05 | 0.04 | -0.12 - 0.02 |
| Beta Time | 0.00 | 0.01 | -0.02 - 0.02 |
| Difference (NT-OLP) | 0.13 | 0.26 | -0.37 - 0.65 |
| MU NT | 0.25 | 0.31 | -0.36 - 0.87 |
| MU OLP | 0.12 | 0.32 | -0.51 - 0.76 |
| P1 (>0) | 0.70 | 0.46 | 0.00 - 1.00 |
| P2 (≥0.2) | 0.39 | 0.49 | 0.00 - 1.00 |
| P3 (≥0.8) | 0.01 | 0.08 | 0.00 - 0.00 |
| Phi | 0.23 | 0.03 | 0.17 - 0.28 |
| Sigma NT | 0.78 | 0.15 | 0.54 - 1.11 |
| Sigma OLP | 0.84 | 0.16 | 0.60 - 1.21 |
| Sigma within | 0.86 | 0.02 | 0.83 - 0.89 |

Abbreviations: GEEE$_{PRE}$ = Generic Rating Scale for Treatment Effects (previous treatment experiences); DESS$_{TAPERING}$ = Discontinuation Emergent Signs and Symptoms Scale (during antidepressant tapering); AR1 = First-Order Autoregressive Error Structure; NT = No Treatment; OLP = Open-Label Placebo; P1 = Posterior Probability of Treatment Effect >0; P2 = Posterior Probability of Treatment Effect ≥0.2; P3 = Posterior Probability of Treatment Effect ≥0.8; CrI = Credible Interval.

Exploratory:

Fixed: Intervention, Time, Prior Experience (GEEE$_{PRE}$), Discontinuation Symptoms During Tapering (DESS$_{TAPERING}$), Maintenance Dose, Duration of Use, Gender

Random: Patients

Autocorrelation: AR1

| Parameter | Mean | SD | CrI |
|---|---|---|---|
| Beta Maintenance Dose | 0.34 | 0.29 | -0.22 - 0.91 |
| Beta Duration of Use | 0.00 | 0.03 | -0.06 - 0.05 |
| Beta Age | 0.00 | 0.01 | -0.02 - 0.02 |
| Beta Gender | 1.02 | 0.36 | 0.31 - 1.75 |
| Beta DESS$_{TAPERING}$ | 3.87 | 0.76 | 2.34 - 5.36 |
| Beta GEEE$_{PRE}$ | -0.02 | 0.04 | -0.09 - 0.06 |
| Beta Time | 0.00 | 0.01 | -0.02 - 0.02 |
| Difference (NT-OLP) | 0.10 | 0.24 | -0.38 - 0.58 |
| MU NT | -1.01 | 0.60 | -2.24 - 0.14 |
| MU OLP | -1.11 | 0.60 | -2.36 - 0.02 |
| P1 (>0) | 0.67 | 0.47 | 0.00 - 1.00 |
| P2 (≥0.2) | 0.34 | 0.47 | 0.00 - 1.00 |
| P3 (≥0.8) | 0.00 | 0.04 | 0.00 - 0.00 |
| Phi | 0.23 | 0.03 | 0.17 - 0.28 |

| | | | |
|---|---|---|---|
| Sigma NT | 0.71 | 0.15 | 0.47 - 1.05 |
| Sigma OLP | 0.79 | 0.15 | 0.55 - 1.14 |
| Sigma within | 0.86 | 0.02 | 0.83 - 0.89 |

Abbreviations: $GEEE_{PRE}$ = Generic Rating Scale for Treatment Effects (previous treatment experiences); $DESS_{TAPERING}$ = Discontinuation Emergent Signs and Symptoms Scale (during antidepressant tapering); AR1 = First-Order Autoregressive Error Structure; NT = No Treatment; OLP = Open-Label Placebo; P1 = Posterior Probability of Treatment Effect >0; P2 = Posterior Probability of Treatment Effect ≥0.2; P3 = Posterior Probability of Treatment Effect ≥0.8; CrI = Credible Interval.

eTable 9. Adverse events

| | Occurrence | Severity[a] | Causality[b] | Occurrence | Severity[a] | Causality[b] | Comparison[c] |
|---|---|---|---|---|---|---|---|
| | *N* (%) [yes/no] | *M* (*SD*) [1-5] | *M* (*SD*) [1-4] | *N* (%) [yes/no] | *M* (*SD*) [1-5] | *M* (*SD*) [1-4] | *Test statistics* |
| **Study visit 2 weeks (t6)** | **OLP (*n* = 12)** | | | **NT (*n* = 13)** | | | |
| *Adverse events* | 3 (25) | 1.00 (0.00) | 3.33 (0.58) | 3 (23) | 1.00 (0.00) | 3.25 (0.35) | *t*(22.17) = 0.09, *p* = .928 |
| **Study visit 4 weeks (t7)** | **OLP (*n* = 13)** | | | **NT (*n* = 12)** | | | |
| *Adverse events* | 3 (23) | 1.00 (0.00) | 3.67 (0.58) | 4 (33) | 1.25 (0.50) | 3.25 (0.50) | *t*(22.13) =-0.55, *p* = .589 |
| **Study visit 6 weeks (t8)** | **OLP (*n* = 12)** | | | **NT (*n* = 13)** | | | |
| *Adverse events* | 3 (25) | 1.0 (0.00) | 3.00 (1.00) | 2 (15) | 1.50 (0.71) | 3.50 (0.71) | *t*(21.48) = 0.58, *p* = .571 |
| **Study visit 8 weeks (t9)** | **OLP (*n* = 13)** | | | **NT (*n* = 12)** | | | |
| *Adverse events* | 0 (0) | - | - | 1 (8) | 1 (-) | 4.00 (-) | *t*(11) = -1.00, *p* = .339 |
| *Total AEs* | 9 | - | - | 10 | - | - | |

*Note.* Results for adverse events related to study participation assessed via clinical interview at biweekly study visits during the N-of-1 trial phase. Adverse events were classified according to the Common Terminology Criteria for Adverse Events (CTCAE).[5]

[a]Severity was rated as grade 1 'mild', grade 2 'moderate', grade 3 'severe', grade 4 'life-threatening', grade 5 'death'.

[b]Causality to study participation was assigned in accordance with the World Health Organization (1 'certain', 2 'probable/likely', 3 'possible', 4 'unlikely', 5 'conditional/unclassified', 6 'unassessable/unclassifiable'). Only adverse events that were related to the study (causality 1-4) were compared.

[c]Occurrence of adverse events was compared for open-label placebo and no treatment periods.

Abbreviations: OLP = Open-Label Placebo; NT = No Treatment; t = Timepoint; n = Number; M = Mean; SD = Standard Deviation; AE = Adverse Event.

eFigure 1. Quality Control Bayesian Analyses – Convergence of the MCMC chain

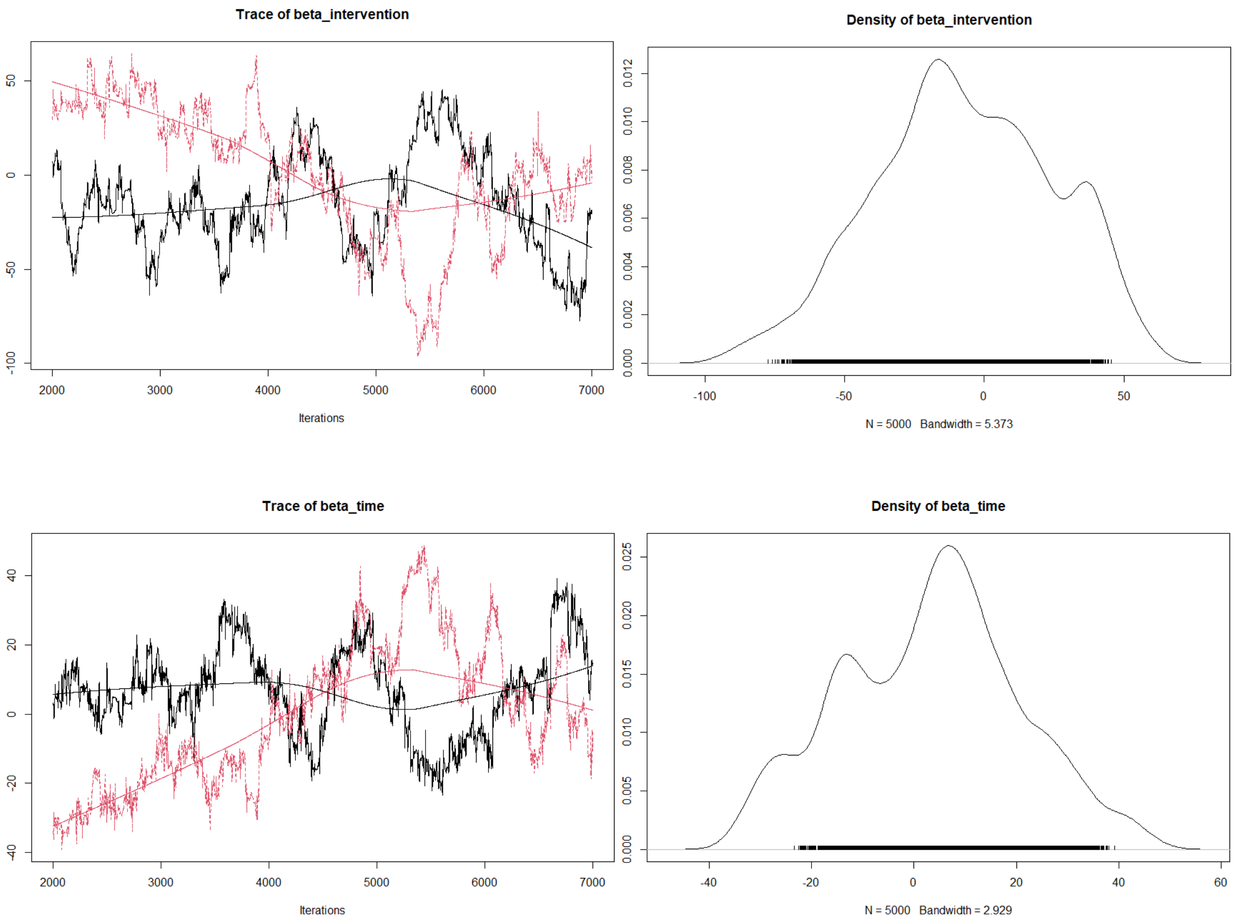


*Note.* Convergence of the Markow-Chain-Monte-Carlo (MCMC) chain was assessed visually via trace plots and density plots for the Bayesian model at individual and population level. Example trace plots and density plots for MCMC chain convergence for the Bayesian model at individual level are presented with data from patient 22 for the primary outcome discontinuation symptoms. The plots of chains for patient 22 are not converged. Similar plots of chains were found for patient 1,3,16 and therefore not considered in the results of the individual analyses in this study.

eMethods 1. Bayesian Model to Estimate the Treatment Effect at Individual Level

```
model
{
  GEEE_act[1] ~ dnorm(mu[1], tau)
  mu[1] <- beta_intercept + beta_intervention * intervention[1] + beta_time * assessment_new[1]

  # AR(1) error structure
  for (i in 2:N) {
    # AR(1) process for GEEE_act
    GEEE_act[i] ~ dnorm(mu[i] + rho * (GEEE_act[i-1] - mu[i-1]), tau_residual)
    mu[i] <- beta_intercept + beta_intervention * intervention[i] + beta_time * assessment_new[i]
  }
  beta_intercept ~ dnorm(0, 0.001)
  beta_intervention ~ dnorm(0, 0.001)
  beta_time ~ dnorm(0, 0.001)
  rho ~ dunif(-1, 1)

  sigma_residual ~ dunif(0.01, 10)
  tau_residual <- 1 / (sigma_residual * sigma_residual)
  sigma_likelihood ~ dunif(0.01, 10)
  tau <- 1 / (sigma_likelihood * sigma_likelihood)

  # Compute mean discontinuation symptom score during OLP and no treatment (NT)
  mu_NT <- beta_intercept + beta_intervention * 0 + beta_time * mean(assessment_new)
  mu_OLP <- beta_intercept + beta_intervention * 1 + beta_time * mean(assessment_new)

  # Difference between mean discontinuation symptom score between OLP and no treatment
  diff <- mu_NT - mu_OLP

  # Probability of clinical meaningfulness
  p1 <- step(diff)          # Probability that the difference is greater than 0
  p2 <- step(diff - 0.2)    # Probability that the difference is greater than 0.2
  p3 <- step(diff - 0.8)    # Probability that the difference is greater than 0.8
}
```

*Note.* The method for estimating probabilities was adapted from the approach described by Stunnenberg et al. (2018).[6] Details of the analysis can be found in the statistical analysis plan: https://clinicaltrials.gov/study/NCT05051995

Abbreviations: N = Number of Observations; NT = No Treatment, OLP = Open-Label Placebo, $GEEE_{ACT}$ = Generic Rating Scale for Treatment Effects (i.e. discontinuation symptoms).

eMethods 2. Bayesian Mixed Model to Estimate the Treatment Effect at Population Level (Basic Model)

```
model
{
 for (i in 1:n_patients) {
  mu_NT[i] ~ dnorm(mu_NT_group, tau_NT)
  mu_OLP[i] ~ dnorm(mu_OLP_group, tau_OLP)
  for (t in 1:N_NT[i]) {
   GEEE_act_NT[i, t] ~ dnorm(mu_NT[i], tau_within)
  }
  for (t in 1:N_OLP[i]) {
   GEEE_act_OLP[i, t] ~ dnorm(mu_OLP[i], tau_within)
  }
 }

 mu_NT_group ~ dnorm(0, 0.001)
 mu_OLP_group ~ dnorm(0, 0.001)

 tau_NT <- 1 / (sigma_NT * sigma_NT)
 sigma_NT ~ dunif(0.01, 10)

 tau_OLP <- 1 / (sigma_OLP * sigma_OLP)
 sigma_OLP ~ dunif(0.01, 10)

 tau_within <- 1 / (sigma_within * sigma_within)
 sigma_within ~ dunif(0.01, 10)

 # Population-level difference in mean discontinuation symptoms during OLP vs no treatment (NT)
 diff <- mu_NT_group - mu_OLP_group

 # Probability of clinical meaningfulness
  p1 <- step(diff)        # Probability that the difference is greater than 0
  p2 <- step(diff - 0.2)   # Probability that the difference is greater than 0.2
  p3 <- step(diff - 0.8)   # Probability that the difference is greater than 0.8
}
```

*Note.* The method for estimating probabilities was adapted from the approach described by Stunnenberg et al. (2018).[6] Details of the analysis can be found in the statistical analysis plan: https://clinicaltrials.gov/study/NCT05051995

Abbreviations: n = Number of Patients, NT = No Treatment, OLP = Open-Label Placebo, $GEEE_{ACT}$ = Generic Rating Scale for Treatment Effects (i.e. discontinuation symptoms).

Supplement
